# Atopy improves survival and decreases risk of brain metastasis in cutaneous melanoma

**DOI:** 10.1101/2024.05.15.24307061

**Authors:** Corey Neff, Mackenzie Price, Gino Cioffi, Zhen Liu, Rabina Walsh, Jill S. Barnholtz-Sloan, Kyle M. Walsh, April K. S. Salama, Carey K. Anders, Peter E. Fecci, Quinn T. Ostrom

**Author notes:** These authors contributed equally to this manuscript and should be considered co-first authors (Corey Neff, Mackenzie Price). **Corresponding Author:** Quinn T. Ostrom, Ph.D., M.P.H. Duke University School of Medicine DUMC Box 3050 Durham, NC 27710.

## Abstract

**Importance:** Development of new therapies in melanoma has increased survival, and as a result more patients are living to develop brain metastasis (BrM). Identifying patients at increased risk of BrM is therefore of significant public health importance.

**Objective:** To determine whether history of atopy is associated with improved survival or reduced incidence of BrM in cutaneous melanoma.

**Design:** A retrospective cohort study conducted from June 2022 to March 2024.

**Setting:** Population-based in states with Surveillance, Epidemiology and End Results (SEER) supported cancer registries.

**Participants:** Individuals (≥65 years) diagnosed with cutaneous melanoma between January 1, 2008 and December 31, 2017 that are participants in traditional Medicare.

**Exposure(s):** Individuals were compared that had history of atopy (allergic rhinitis, atopic dermatitis, asthma, and/or allergic/atopic conjunctivitis) diagnosed prior to melanoma diagnosis, ascertained using ICD-9 or ICD-10 codes in Medicare claims.

**Main Outcome(s) and Measure(s):** Primary endpoints were diagnosis with a BrM or death during the follow-up period. Associations between atopy and endpoints were assessed using cox proportional hazards models to estimate hazard ratios (HR) and p-values.

**Results:** A total of 29,956 cutaneous melanoma cases were identified (median age 76, 60% male and 97% non-Hispanic White). Overall, 7.1% developed BrM during follow up. Among the 35% that had history of atopy, the most common condition was atopic dermatitis (19%). After adjustment for demographic and prognostic factors, atopy was associated with a 16% decrease in death (HR=0.84 [95%CI:0.80-0.87], p_FDR_<0.001). Among those with non-metastatic disease at time of diagnosis, atopy conferred a 15% decrease in cumulative incidence BrM (HR=0.85 [95%CI: 0.76-0.94], p_FDR_=0.006), with a 25% decrease associated with atopic dermatitis (HR=0.75 [95%CI:0.65-0.86], p_FDR_<0.001). Among those with metastatic disease at diagnosis (any metastatic site), only those who received immune checkpoint inhibitors had a survival benefit associated with atopy (HR=0.31, [95%CI:0.15-0.64], p=0.001 vs HR=1.41, [95%CI:0.87-2.27], p=0.165).

**Conclusions and Relevance:** Atopy, particularly atopic dermatitis, was significantly associated with improved survival and decreased incidence of BrM. The improved survival associated with these conditions in the context of immunotherapy suggests that these conditions in the elderly may identify those with more robust immune function that may be more responsive to treatment.

**Key Points:** **Question:** Does atopy affect outcomes in cutaneous melanoma?

**Findings:** In a retrospective cohort study, elderly individuals with prior diagnosis of atopy had significant improved overall survival and decreased incidence of brain metastasis as compared to individuals without atopy.

**Meaning:** History of atopy may identify a subgroup within melanoma cases that has improved response to treatment and a more robust immune system which decreases risk of metastasis to the brain.

## INTRODUCTION

Allergic/atopic diseases are among the most common medical conditions in the United States (US), and risk is modified by genetics and environment. These conditions result in a Th2-skewed immune response, with elevated IgE and propensity for hypersensitivity.^1, 2^ The relationship between atopy and cancer is complex, reflecting both the pro and anti-tumorigenic effects of chronic inflammation and immune activation, respectively.^3-7^ Melanoma is one of the most immune-infiltrated cancers and is a major success story in cancer immunotherapy. These tumors occur more commonly in t immunosuppressed persons,^8^ and are frequently enriched with infiltrating immune cells. Prior studies examining associations between melanoma risk and atopy have produced mixed results.^3-7, 9^ Substantially less research has evaluated the relationship between pre-existing atopy and melanoma outcomes.

Secondary malignancies of the brain, or brain metastases (BrM), are the most common type of tumor in the central nervous system (CNS), occurring with a frequency several times that of primary brain tumors (PBT). Treatment advances that improve control of local disease and extend overall survival (OS) have likely led to a concomitant increase in the proportion of individuals developing BrM, despite continued decreases in cancer incidence.^10-12^ As a result, frequency of BrM is on the rise,^13^ particularly among individuals diagnosed with melanoma, lung, and breast cancer. Prior work found 1.2% of melanoma patients present with BrM (synchronous BrM, sBrM), while ∼7% develop BrM during treatment.^14-16^ Several factors have been associated with an increased risk of BrM, including: younger age, Breslow thickness, higher stage at diagnosis, and scalp location.^17-20^

Given the significant morbidity and mortality associated with BrM, identifying populations at elevated risk is of significant clinical importance. Atopy is consistently associated with decreased risk of PBT,^21-24^ and has also been associated with improved OS,^25^ presumably due to increased CNS immune surveillance. Whether insights from PBT also extend to BrM remains unknown, but the impact atopy has on risk of developing BrM merits investigation. In this analysis, we aimed to assess the relationship between pre-existing atopy, risk of BrM, and OS among patients with cutaneous malignant melanoma.

## METHODS

This project was approved as an exempt protocol by the Duke University School of Medicine Institutional Review Board. The National Cancer Institute’s Surveillance, Epidemiology, and End Results (SEER) Program includes specially funded population-based cancer registries that collect data on newly diagnosed cancers within catchment area (∼24% of the US population^26^). For individuals that are enrolled in Medicare, existing SEER case data can be linked to Medicare claims.

International Classification of Diseases, Oncology 3^rd^ edition (ICD-O-3) codes were used to identify cutaneous melanoma (ICD-O-3 morphology codes 8720/3-8790/3 and topographic codes C44.0-C44.9) cases for diagnoses made from January 1, 2008 to December 31, 2017 with corresponding Medicare claims through December 31, 2019. Cases were included when ≥65 years and melanoma was the individual’s only lifetime cancer diagnosis. Cases were excluded if they were: 1) qualified for Medicare for a reason other than age; 2) enrolled in a Health Maintenance Organization during a period six months prior to twelve months after diagnosis; 3) not continuously enrolled in Part A & B for 6 months prior to diagnosis through the end of follow-up; or 4) missing race/ethnicity, sex, stage, or follow-up. Total exclusions represented 51.4% of cases (**eFigure 1**).

**Figure 1.**
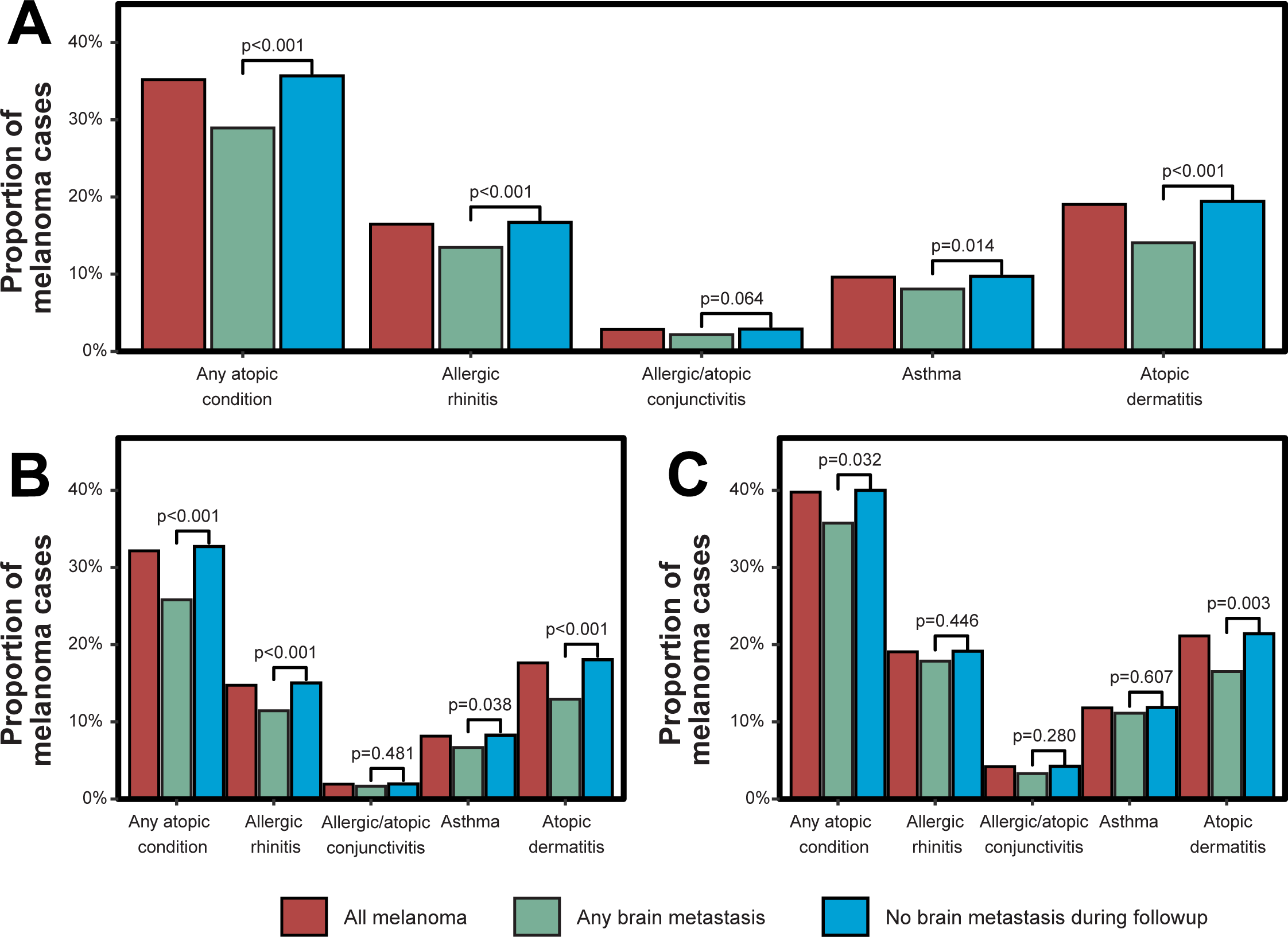
Frequency of atopic conditions at time of melanoma diagnosis by condition A) over-all, B) in males only, and C) in females only.

Prior atopy was identified by ICD-9/10 diagnosis codes **(eTable 1)** and each condition was defined by ≥2 claims of the same code ≥6 months before melanoma diagnosis. A SEER-provided variable was used to identify sBrM). Additional BrM, including those that were discovered after initial diagnosis (asynchronous BrM, aBrM), were identified using the Medicare Provider Analysis and Review (MEDPAR), National Claims History (NCH), and Outpatient (OUTPAT) claims files. BrM was defined as an ICD-9/ICD-10for BrM and ≥1 procedure code for a brain/head diagnostic test within 60 days of the ICD-9/ICD-10 code **(eTable 2)**. BrM were defined as synchronous if within 30 days of diagnosis and were otherwise classified as asynchronous. Stage was defined using SEER summary stage. Comorbidity score was estimated using the SEER-Medicare SAS comorbidity macro^27^ based on ICD-9/ICD-10 codes with a modified Charlson weight. Cancer treatment, including surgery, radiation, chemotherapy, or immune checkpoint inhibitor use were identified using SEER variables and claims **(eTable 2)**. Radiation, chemotherapy, and ICI were defined by ≥2 claims for the same code separated by at ≥1 week. Tumor size was defined using Collaborative Stage (CS) Tumor size, and American Joint Committee on Cancer (AJCC) T staging. Surgical size was defined using CPT codes **(eTable 2)**. Breslow depth was defined using CS Site-Specific Factor (SSF) 1 and ulceration was defined using CS SSF 2. Survival and vital status were defined using the SEER-provided variables. Missing/invalid values were imputed using date of death or last follow-up, when available. Cumulative incidence of BrM was defined as the time between diagnosis and date of aBrM or date of last follow-up, with those that died before development of BrM being right-censored.

The odds of receiving treatment based on atopy status were assessed using multivariable logistic regression models, adjusting for demographics, stage at diagnosis, treatment, and comorbidity score. Cox proportional hazards models were used to assess the relationship between OS in all melanoma cases and hazard of developing BrM among those with localized or regional disease at diagnosis. Competing risks analyses were performed using R package cmprsk to estimate the subdistribution hazard of developing a BrM while controlling for the competing risk of death (all cause).^28^ In order to account for potential false positives due to multiple testing, p-values were adjusted in multivariable models (where noted) using the Benjamini-Hochberg method, and significance was assessed by controlling the false discovery rate (FDR) at α=0.05. All analyses were conducted using R version 4.1.3.

## RESULTS

After all exclusions there were 29,956 cases of melanoma, with median age at diagnosis of 76 years. Overall, 60% were male, and 97% were non-Hispanic White (NHW). Among these individuals, 7% developed BrM during the follow up period **(Table 1)**. Males developed BrM more frequently (8.1% vs 5.5%, p<0.001), and males with BrM were more likely to have sBrM (20.5% vs 17.1%), but this difference was not statistically significant (**Figure 1**). While melanoma is much more common in NHW, the proportion developing BrM was significantly higher in all other racial/ethnic groups (6.9% in NHW, 11.0% in Hispanics, and 19% in non-Hispanic Blacks [NHB], p<0.001) (**Figure 1**, **Table 1**). In those with BrM, NHW were more likely to have sBrM, but this difference was not statistically significant. There was no significant difference in age at diagnosis by BrM. Known prognostic factors varied significantly between those with and without BrM, and there was substantial correlation among prognostic factors (**eFigure 2**). In general, those with BrM had larger primary tumors, deeper extension, more extensive surgery, and were more likely to have ulceration. The most common primary site among those with BrM was “other or site unspecified” (28%), which includes melanoma of unknown primary.

**Figure 2.**
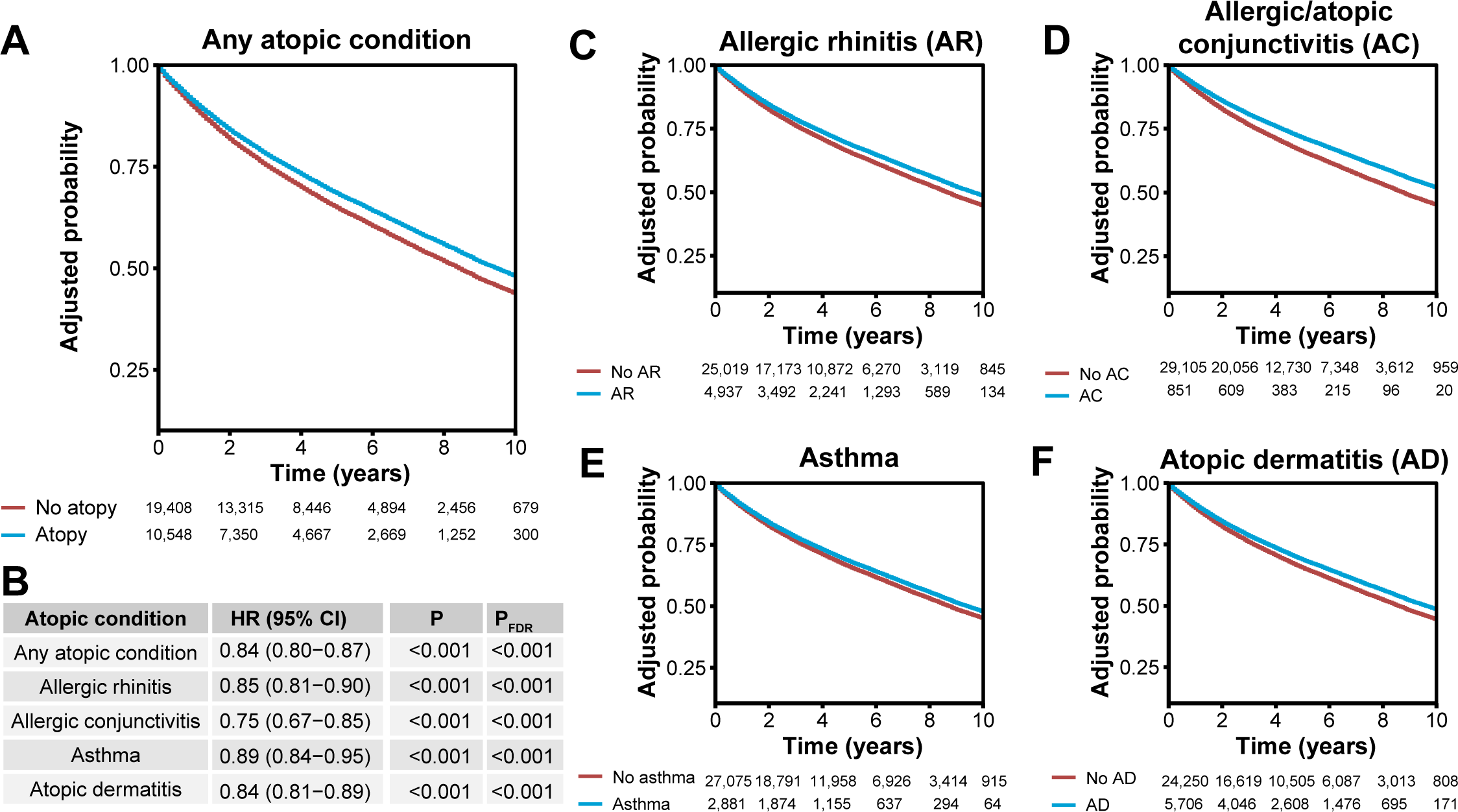
Adjusted^a^ probability of survival for melanoma by prior history of A) any atopic con-dition, and B) corresponding hazard ratios, 95% CIs and p-values, and adjusted probability of survival by prior history of C) allergic rhinitis, D) allergic/atopic conjunctivitis, E) asthma, and F) atopic dermatitis. a. Adjusted for age at diagnosis, sex, race/ethnicity, comorbidity score, stage at diagnosis, primary site, tumor size, depth of invasion, presence of ulceration, and treatment receipt (including resection, radiation, and chemotherapy)

**Table 1.** Characteristics of melanoma cases in SEER-Medicare by brain metastasis status.

| Characteristic | Overall,<br>N = 29,956 | Extracranial<br>disease only<sup>a</sup><br>(never developed<br>brain metastasis)<br>N = 27,838 | Any brain<br>metastasis<br>N = 2,118 | p-value<sup>b</sup> |
| --- | --- | --- | --- | --- |
| **Median survival months (95% CI)** | 101 (99-104) | 111 (108-114) | 21 (20-23) | **<0.001** |
| **Male (%)** | 17,917 (60%) | 16,465 (59%) | 1,452 (69%) | **<0.001** |
| **Median age at diagnosis (IQR)** | 76 (70, 82) | 76 (70, 83) | 75 (70, 81) | **<0.001** |
| **Race/Ethnicity** |  |  |  | **<0.001** |
| Non-Hispanic White | 29,180 (97%) | 27,159 (98%) | 2,021 (95%) |  |
| Non-Hispanic Black | 112 (0.4%) | 91 (0.3%) | 21 (1.0%) |  |
| Non-Hispanic Other | 160 (0.5%) | 138 (0.5%) | 22 (1.0%) |  |
| Hispanic | 504 (1.7%) | 450 (1.6%) | 54 (2.5%) |  |
| **Stage at diagnosis** |  |  |  | **<0.001** |
| Localized | 24,906 (83%) | 24,133 (87%) | 773 (36%) |  |
| Regional | 1,848 (6.2%) | 1,012 (3.6%) | 836 (39%) |  |
| Distant site involved | 3,202 (11%) | 2,693 (9.7%) | 509 (24%) |  |
| **Tumor size / AJCC T staging** |  |  |  | **<0.001** |
| < 1.0 mm (T1) | 9,951 (33%) | 9,780 (35%) | 171 (8.1%) |  |
| >1-2 mm (T2) | 2,505 (8.4%) | 2,337 (8.4%) | 168 (7.9%) |  |
| >2-4 mm (T3) | 2,438 (8.1%) | 1,971 (7.1%) | 467 (22%) |  |
| >4 mm (T4) | 3,372 (11%) | 3,006 (11%) | 366 (17%) |  |
| Unknown | 11,690 (39%) | 10,744 (39%) | 946 (45%) |  |
| **Size of surgical excision** |  |  |  | **<0.001** |
| <3.1 cm | 13,245 (44%) | 12,587 (45%) | 658 (31%) |  |
| >3.0 cm | 8,910 (30%) | 8,440 (30%) | 470 (22%) |  |
| None/Unknown | 7,801 (26%) | 6,811 (24%) | 990 (47%) |  |
| **Primary Site** |  |  |  | **<0.001** |
| Head and neck | 8,124 (27%) | 7,785 (28%) | 339 (16%) |  |
| Lower limb and hip | 4,528 (15%) | 4,287 (15%) | 241 (11%) |  |
| Upper limb and shoulder | 8,124 (27%) | 7,785 (28%) | 339 (16%) |  |
| Trunk | 7,917 (26%) | 7,489 (27%) | 428 (20%) |  |
| Other or site unspecified<sup>c</sup> | 1,414 (4.7%) | 811 (2.9%) | 603 (28%) |  |
| **Breslow depth of invasion** |  |  |  | **<0.001** |
| < 1.0 mm | 13,033 (44%) | 12,603 (45%) | 430 (20%) |  |
| >1-2 mm | 3,331 (11%) | 3,154 (11%) | 177 (8.4%) |  |
| >2-4 mm | 2,342 (7.8%) | 2,076 (7.5%) | 266 (13%) |  |
| >4 mm | 2,266 (7.6%) | 1,926 (6.9%) | 340 (16%) |  |
| Unknown | 8,984 (30%) | 8,079 (29%) | 905 (43%) |  |
| **Ulceration** |  |  |  | **<0.001** |
| Present | 3,822 (13%) | 3,283 (12%) | 539 (25%) |  |
| Not present | 17,071 (57%) | 16,515 (59%) | 556 (26%) |  |
| Unknown | 9,063 (30%) | 8,040 (29%) | 1,023 (48%) |  |
| **Received surgery for primary tumor (%)** | 24,797 (83%) | 23,412 (84%) | 1,385 (65%) | **<0.001** |
| **Received radiation treatment (%)** | 1,576 (5.3%) | 783 (2.8%) | 793 (37%) | **<0.001** |
| **Received chemotherapy (%)** | 3,282 (11%) | 2,416 (8.7%) | 866 (41%) | **<0.001** |
| **Received checkpoint immunotherapy (%)** | 2,162 (7.2%) | 1,449 (5.2%) | 713 (34%) | **<0.001** |
| **Comorbidity Score** |  |  |  | 0.546 |
| 0 | 16,227 (54%) | 15,088 (54%) | 1,139 (54%) |  |
| 1 | 4,644 (16%) | 4,317 (16%) | 327 (15%) |  |
| 2 | 4,656 (16%) | 4,305 (15%) | 351 (17%) |  |
| 3+ | 4,429 (15%) | 4,128 (15%) | 301 (14%) |  |
| **Brain metastasis timing** |  |  |  | **<0.001** |
| Synchronous | -- | -- | 408 (19%) |  |
| Asynchronous | -- | -- | 1,710 (81%) |  |
| **Any atopic condition (%)** | 10,548 (35%) | 9,935 (36%) | 613 (29%) | **<0.001** |
| Allergic rhinitis | 4,937 (16%) | 4,652 (17%) | 285 (13%) | **<0.001** |
| Atopic dermatitis | 5,706 (19%) | 5,408 (19%) | 298 (14%) | **<0.001** |
| Asthma | 2,881 (9.6%) | 2,710 (9.7%) | 171 (8.1%) | **0.014** |
| Allergic/atopic conjunctivitis | 851 (2.8%) | 805 (2.9%) | 46 (2.2%) | 0.055 |
| **Number of atopic conditions** |  |  |  |  |
| 0 | 19,673 (66%) | 18,153 (65%) | 1,520 (72%) | **<0.001** |
| 1 | 7,502 (25%) | 7,036 (25%) | 466 (22%) |  |
| 2 | 2,321 (7.7%) | 2,213 (7.9%) | 108 (5.1%) |  |
| 3+ | 460 (1.5%) | 436 (1.6%) | 24 (1.1%) |  |

<sup>a</sup> Includes individuals with no metastasis as well as extracranial metastasis only.
<sup>b</sup> p-value corresponding to either log-rank test for median survival, two-tailed t-test for age at diagnosis, or $\chi^2$ test of independence for all others.
<sup>c</sup> Includes metastatic melanoma with unknown primary.

Among all melanoma cases, 35% had prior atopy and the most common atopic condition was atopic dermatitis (19%) (**Table 1**). Individuals with BrM were less likely to have atopy (29%) as compared to individuals without (36%) (**Figure 1**). Frequency of all atopic conditions was higher in females **(Figure 1**), and there was no significant difference by race/ethnicity (**eTable 3**). Individuals with atopy were more likely to have received chemotherapy treatment (odds ratio [OR]=1.15, p_FDR_=0.01) and radiation treatment (OR=1.16, p_FDR_=0.047) as compared to those without atopy (**eFigure 4**). Allergic rhinitis was significantly associated with increased chemotherapy use (OR=1.20, p_FDR_=0.016) and increased use of ICI (OR=1.15, p_FDR_=0.08).

Median OS was 101 months. OS varied significantly by sex (96 months in males vs 109 months in females, p<0.001). Known prognostic factors for melanoma were significantly associated with OS in univariable models (**eTable 4**). Atopy was associated with a significant survival benefit (HR=0.84, p_FDR_<0.001) in all melanoma cases after adjustment for demographic and prognostic factors, and a ∼15% reduction in hazard of death was consistent across atopic diseases (e**Figure 5**). Individuals with ≥2 atopic conditions had a ∼16% significant decrease in hazard of death (HR=0.84) compared to those without atopy, in comparison to a ∼11% decrease in those with only one atopic condition (**eTable 5**). Effect of atopy varied by age at diagnosis, with younger individuals with atopy having a poorer outcome and older individuals experiencing a stronger protective association (**eFigure 6**). In an interaction analysis, there was a non-significant statistical interaction between age and atopy on OS (p=0.060). Individuals with atopy were more likely to be diagnosed with localized disease (p<0.001, **eTable 3**). Among those with BrM only, there was no OS benefit associated with any atopic condition (**eFigure 7**), and no significant OS benefit in the sBrM or aBrM subsets (**eFigure 8-9**).

Among those with localized or regional disease at diagnosis who developed an aBrM, median time was 19.2 months. Known prognostic factors for melanoma were associated with cumulative incidence of aBrM. Prior atopy was associated with a significant decrease in risk for aBrM (HR=0.85, p_FDR_=0.006), but this was no lower significant after adjustment for competing risk of death (**eFigure 10**, **Figure 3**). When conditions were evaluated individually, only atopic dermatitis was associated with decreased risk of aBrM (HR=0.75. p_FDR_<0.001), which remain significant after adjustment for competing risk of death (subdistribution HR=0.81, p_FDR_=0.022). Increased number of atopic conditions was associated with decreased risk of aBrM, but there was not a statistically significant dose-response (**eTable 5**).

**Figure 3.**
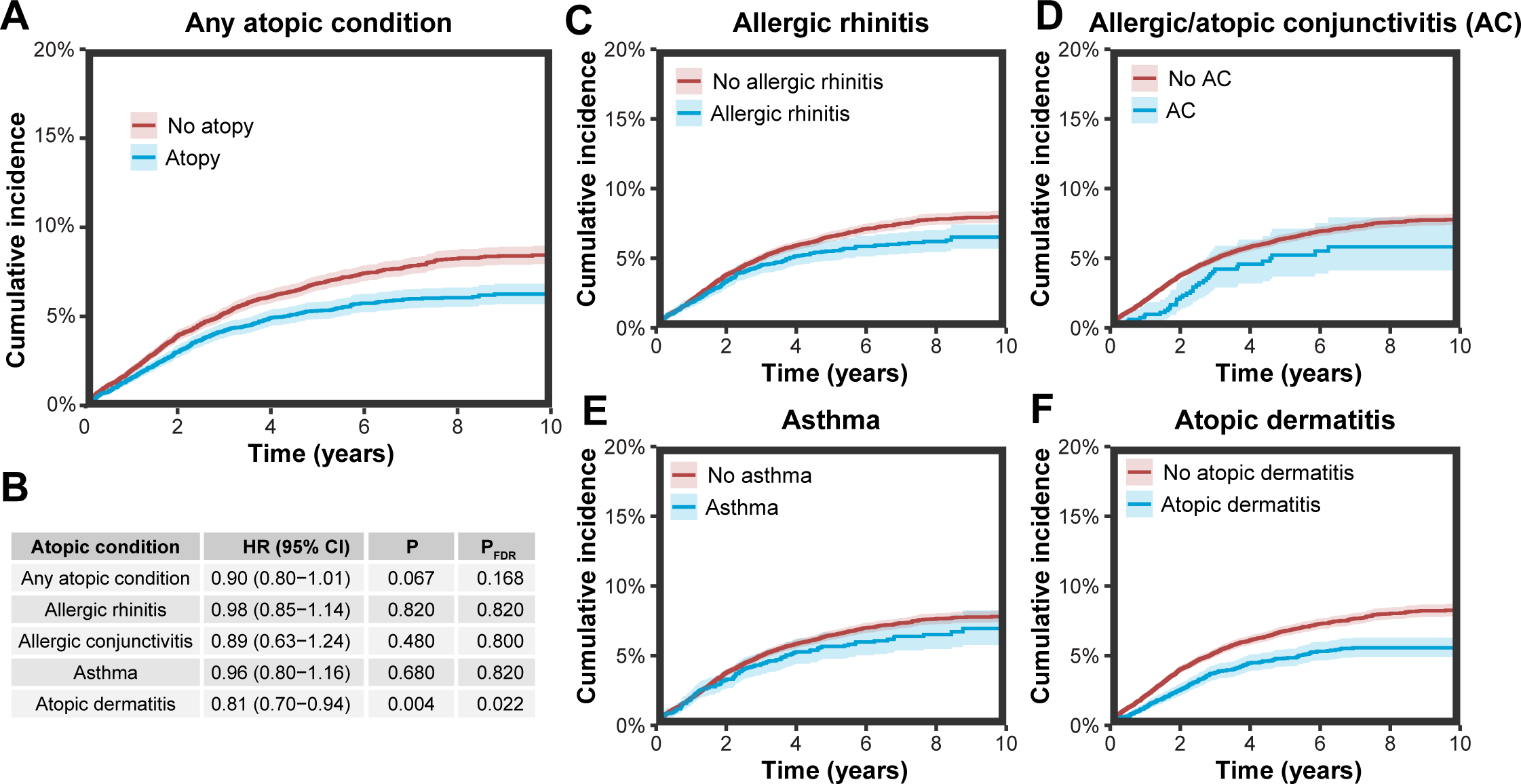
Adjusted^a^ subdistribution hazard of developing an asynchronous brain metastasis after adjustment for competing risk of death with 95% CI by prior history of A) any atopic con-dition, and B) corresponding hazard ratios, 95% CIs and p-values, and adjusted probability of survival by prior history of C) allergic rhinitis, D) allergic/atopic conjunctivitis, E) asthma, and F) atopic dermatitis. a. Adjusted for age at diagnosis, sex, race/ethnicity, comorbidity score, stage at diagnosis, primary site, tumor size, depth of invasion, presence of ulceration, and treatment receipt (including resection, radiation, and chemotherapy)

In order to account for screening bias that may have resulted in earlier diagnosis among those with atopic skin disease, sensitivity analyses were completed in individuals with regional disease at diagnosis only (N=1,848). Atopic dermatitis was still associated with improved OS, though the association was attenuated and non-significant (HR=0.91, p=0.252). Atopic dermatitis remained protective against aBrM, though the association was attenuated and non-significant (HR=0.85, p=0.365) (**eTable 5**). Among those with distant spread at diagnosis without BrM, atopic dermatitis was significantly associated with improved OS (HR=0.64, p=0.018). Among those with distant disease at diagnosis but no BrM, atopic dermatitis was associated with a significant 63% decrease in cumulative incidence of aBrM (HR=0.37, p=0.006).

In the subset of individuals with distant disease at diagnosis diagnosed after approval of ICI in advanced melanoma (2012-2017), atopy was associated with lower frequency of sBrM, though the difference was non-significant (5.3% vs 15.8%, p=0.086). Only 2.7% of those with atopic dermatitis and metastatic disease had sBrM (p=0.521). Atopy and atopic dermatitis were associated with improved OS those with distant disease at diagnosis (any metastatic site) that received ICI but no improvement in those that did not receive ICI (**eTable 5**).

## DISCUSSION

The results of this study demonstrate that pre-existing atopy in melanoma is associated with both improved OS and decreased risk of BrM. Among individual atopic conditions, the strongest effects were identified for atopic dermatitis. While others have identified mixed associations between risk of melanoma and atopy,^3-7^ to our knowledge, this is the first assessment of the effect of pre-existing exaggerated immune response on melanoma outcomes.

Prevalence of atopy was 34%, and prevalence of specific conditions is similar to previously published estimates in the elderly.^29^ Cumulative incidence of BrM was 7.6%, which is consistent with prior estimates.^15, 16^ Risk for BrM varied significantly between demographic groups, with highest frequency of BrM in males and NHB. This risk was multiplicative, with ∼25% of NHB males developing BrM. Highest frequency of atopy was observed in females, but there was no difference in frequency of atopy by race/ethnicity. Prevalence of atopy is higher in females, which may combine with additional behavioral factors (*e.g.*, sunscreen use, frequency of dermatologic screenings) to which may contribute to lower incidence of melanoma among women.

The most consistent atopic disease association identified in this analysis was with atopic dermatitis, an outwardly visible condition that frequently co-occurs with IgE-mediated respiratory conditions including allergic rhinitis. A relationship with the immune milieu of the CNS is plausible, as prior analyses have shown that atopic dermatitis increases lifetime risk of dementia and stroke, and may reduce risk of glioma and meningioma.^24, 30-32^ Melanoma and atopic dermatitis are both dermatologic conditions, and it is likely that someone with existing atopic dermatitis may be more likely to receive a melanoma diagnosis at an earlier stage than someone without a dermatologic condition. Prior analyses based on occupational examinations for skin cancers (screened regardless of dermatologic disease status) have found significantly lower incidence among those with atopic conditions,^6^ suggesting that this association is not solely the result of screening bias. Furthermore, when our analyses were limited to those with more advanced melanoma at diagnosis (excluding early stage diagnoses that may be more common in those under regular dermatologist care), the protective effects of atopy were consistent with those in the full population.

The potential protective effect of atopic disease on cancer progression observed in this analysis has previously been observed in other cancer types, including colorectal cancer and glioma.^33,25^ There are several mechanisms through which atopic disease could inhibit cancer progression. Allergic reactions recruit eosinophils to the affected area and also increase the proportion of these cells in overall circulating leukocyte populations.^34, 35^ The anticancer effect of eosinophils has been previously observed,^36, 37^ including in melanoma, where both increased circulating and intratumoral eosinophils predicted improved responses to immunotherapy.^38-41^ The mechanism through which eosinophils exert anti-tumor functions is not well understood. In a murine melanoma model, a prior study found that IL-33 stimulated recruitment of eosinophils to the lungs and significantly delayed development of pulmonary metastasis.^42^ Another potential mechanism through which eosinophils are hypothesized to increase anti-cancer immune responses is by modifying the activity of effector T cells. Eosinophils have been observed to stimulate both primed and naïve T cells, to activate T cell proliferation, and in some circumstances, to present antigen to T cells in draining lymph nodes.^43^ Therapies for atopic conditions may also contribute to the effects observed here. These drugs include steroids and other anti-inflammatories, disease-modifying antirheumatic drugs (DMARD), and anti-IgEs. Some DMARDs, such as methotrexate, are used as chemotherapies for non-melanoma cancers, and there has been some suggestion that use of these may increase risk of melanoma.^44^ Due to the age-related eligibility for Medicare, we are not able to assess the association between lifetime use of these drugs and effect on melanoma risk or outcomes.

Individuals <75 years old did not experience a significant survival benefit from atopy, and individuals 65-69 were significantly less likely to be classified as having atopy (**eTable 3**), suggesting some possible age-related misclassification. Younger individuals will have fewer years of available Medicare claims, and as a result, they may be inaccurately classified as not having atopy if they did not seek medical care after enrolling in Medicare. Immune function gradually declines with increasing age, which often leads to declines in allergic sensitization.^45, 46^ While the clinical manifestations of many atopic conditions present in adulthood decline with age in the elderly, most notably that of allergic asthma, serum IgE often remains elevated and has been shown to decline least in the setting of atopic dermatitis.^46^ The presence of atopic dermatitis in elderly individuals may be a clinical biomarker of decreased immunosenescence and could help to distinguish individuals with more robust immune function than their age-matched peers. A subset analysis conducted only in those with distant disease at diagnosis that received ICI identified a significant survival benefit associated with both atopy, and atopic dermatitis in particular, which was not observed in those with distant disease that did not receive ICI. This suggests that presence of atopy may serve as a marker of a more robust immune system which may be more primed to stimulation by immunotherapy.

While SEER-Medicare data is a powerful resource for research, there are several important limitations. While the vast majority of those ≥65 in the US are entitled to Medicare, there are several factors that affect whether claims are reported. Individuals in the workforce may retain employer-sponsored insurance as primary insurer and Medicare as secondary. In this case, claims would first be billed to the primary insurance. Many individuals select Medicare Advantage (MA) coverage through private insurance companies, which is paid for by Medicare but does not result in claims directly to CMS. In both of these cases, claims for services will be unrepresented in these data. While there are no substantial differences between those choosing traditional Medicare as opposed to MA,^47^ there may be significant differences between those with private insurance, MA, and those with traditional Medicare that would limit the generalizability of our findings. All individuals included in this analysis are ≥65 years, meaning that younger individuals are excluded. The median age of first diagnosis with melanoma is ∼66,^48^ and as a result this should capture >50% of the population with melanoma. Care must be taken when interpreting the use of claims data for metastasis research.^49^ Use of ICD-9/ 10 to identify patients with aBrM is imperfect. While SEER-Medicare contains nearly all claims billed through Medicare, previous research has shown that the information contained in the claims themselves is not always reliable or valid.^50-53^ Threats to the validity of claims may include under/over-diagnosing, patient compliance with billed treatments, and inconsistent use of billing codes, among others. Prior evaluations have demonstrated that use of tumor-specific algorithms have ∼55% sensitivity and 85% specificity for lung cancer, suggesting that individuals with metastasis will be missed but those assigned as having metastasis are likely correctly identified. These algorithms will result in the underestimation of total aBrM.^54^ Classification of atopy with claims data further requires that atopy was severe enough to seek medical care, and as a result, individuals with more mild conditions may be incorrectly classified as not having atopy.

## CONCLUSION

Atopy is associated with moderate improvements in OS in melanoma and with a decreased hazard of developing melanoma BrM in the US Medicare population. These findings suggest that hyperimmunity, for which atopy may be a surrogate, may increase OS and decrease frequency of BrM. Further research is necessary to determine the biological mechanisms through which atopy and other immune hypersensitivities affect prognosis, as well as the extent to which the protective effects of atopy may explain sex differences in risk of BrM.

## Data Availability

All data used in the present study are available upon request and approval of the SEER-Medicare linkage data.

## Acknowledgements

This study used the linked SEER-Medicare database. The interpretation and reporting of these data are the sole responsibility of the authors. The authors acknowledge the efforts of the National Cancer Institute; Information Management Services (IMS), Inc.; and the Surveillance, Epidemiology, and End Results (SEER) Program tumor registries in the creation of the SEER-Medicare database. The collection of cancer incidence data used in this study was supported by the California Department of Public Health pursuant to California Health and Safety Code Section 103885; Centers for Disease Control and Prevention’s (CDC) National Program of Cancer Registries, under cooperative agreement 1NU58DP007156; the National Cancer Institute’s Surveillance, Epidemiology and End Results Program under contract HHSN261201800032I awarded to the University of California, San Francisco, contract HHSN261201800015I awarded to the University of Southern California, and contract HHSN261201800009I awarded to the Public Health Institute. The ideas and opinions expressed herein are those of the author(s) and do not necessarily reflect the opinions of the State of California, Department of Public Health, the National Cancer Institute, and the Centers for Disease Control and Prevention or their Contractors and Subcontractors.

