## Supplemental Tables and Figures for "Atopy improves survival and decreases risk of brain metastasis in cutaneous melanoma"

**eTable 1. Classification of atopic conditions**

| Condition | ICD-10-CM | ICD-9-CM |
|---|---|---|
| **Atopic Conditions** | | |
| Allergic rhinitis | J30.X | 477.X |
| Asthma | J45.X | 493.X, 975.7 |
| Atopic dermatitis | B00.0, H01.13X, H60.54X, L01.1, L20.8, L20.9 | 373.31, 540.X, 691.8, 692.0, 692.1, 692.2, 692.3, 692.4, 692.5, 692.6, 692.84, 692.89, 692.9, 692.9 |
| Allergic/atopic conjunctivitis | H10.1, H10.45 | 372.05, 372.14 |

**eTable 2. Codes used to determine treatment pattern**

| Treatment Type | Code Classification | Code |
|---|---|---|
| Diagnostic Imaging (cranial only) | CPT | 70450-70470, 70551-70553, 78607-78608 |
| Radiation Therapy | ICD-9-CM | 92.23–92.24, 92.30–92.33, and 92.39. |
| | CPT | 77261-77263, 77280, 77285, 77290, 77295, 77299–77301, 77305, 77310, 77315, 77321, 77332-77334, 77336-77337, 77370-77372, 77399, 77402-77414, 77416, 77418-77420, 77425, 77427, 77430, 77432, and 0073T. |
| | HCPCS | G0173-G0174, G0242-G0243, G0251, and G0338-G0340. |
| Stereotactic Radiosurgery | ICD-9-CM | Cranial: 92.30–92.33 and 92.39. |
| | CPT | Cranial: 61793, 61796-61800, 77371-77372, and 77432. |
| | HCPCS | Cranial: G0173, G0242, G0243, G0251, G0338, G0339, and G0340. |
| Surgical Size | CPT | <3.1 cm: 11400, 11401, 11402, 11403, 11420, 11421, 11422, 11423, 11440, 11441, 11442, 11443, 11600, 11601, 11602, 11603, 11620, 11621, 11622, 11623, 11640, 11641, 11642, 11643<br>>3.0 cm: 11404, 11406, 11424, 11426, 11444, 11446, 11604, 11606, 11624, 11626, 11644, 11646 |
| **Immune Checkpoint Inhibitors** | | |
| **Treatment Type** | **Code Classification** | **Code** |
| Pembrolizumab | HCPCS | C9027, J9271 |
| Necitumumab | HCPCS | C9475, J9295 |
| Nivolumab | HCPCS | C9453, J9299 |
| Ipilimumab | HCPCS | C9284, J9228 |
| Atezolizumab | HCPCS | C9483, J9022 |
| Ramucirumab | HCPCS | C9025, J9308 |
| **Chemotherapies** | | |
| Carboplatin | HCPCS | J9045 |
| | NDC | 00015-3210, 00015-3211, 00015-3212, 00015-3213, 00015-3214, 00015-3215, 00015-3216, 61703-0360, 00703-3249, 47335-0150, 47335-0151, 50742-0447, 50742-0448, 66758-0047, 68083-0190, 68083-0191, 68083-0192, 68083-0193, 71288-0100, 00703-4239, 00703-4244, 00703-4246, 57277-0105, 57277-0106, 67457-0491, 67457-0492, 67457-0493, 67457-0494, 67457-0608, 55150-0386, 16729-0295, 69448-0005, 00703-4248, 61703-0339, 63323-0172, 25021-0202, 47335-0284, 47781-0603, 47781-0604, 47781-0605, 47781-0606, 50742-0445, 50742-0446, 57277-0107 |
| Cisplatin | HCPCS | C9418, J9060, J9062 |
| | NDC | 44567-0530, 00015-3070, 00015-3072, 00703-5747, 00703-5748, 16729-0288, 44567-0509, 44567-0510, 44567-0511, 63323-0103, 68001-0283, 68083-0162, 68083-0163, 67457-0424, 67457-0425, 70860-0206, 00069-0081, 00069-0084, 47781-0609, 47781-0610, 61126-0003, 61126-0004 |
| Dacarbazine | HCPCS | C9423, J9130, J9140 |
| | NDC | 00703-5075, 63323-0127, 63323-0128, 61703-0327 |
| Docetaxel | HCPCS | J9170, J9171 |
| | NDC | 70121-1221, 70121-1222, 70121-1223, 43066-0001, 43066-0006, 43066-0010, 00069-9141, 00069-9142, 00075-8001, 00075-8005, 00409-0366, 00409-0367, 00955-1020, 00955-1021, 00955-1022, 25021-0222, 43598-0258, 43598-0610, 43598-0611, 45963-0765, 63739-0932, 63739-0971, 66758-0050, 66758-0950, 25021-0245, 50742-0431, 50742-0463, 00143-9204, 00143-9205, 43598-0389, 47335-0323, 47335-0895, 47335-0939, 72485-0216, 72485-0215, 72485-0214, 71288-0143, 71288-0144, 71288-0150, 71288-0151, 00409-7870, 00409-0365, 00409- |

| | | |
|---|---|---|
| | | 1732, 00409-4235, 00409-5068, 55150-0378, 55150-0379, 55150-0380, 68083-0401, 68083-0400, 68083-0399, 00409-0201, 00409-0368, 16729-0231, 16729-0267, 43598-0259, 45963-0734, 47335-0285, 67457-0533, 67457-0781, 69097-0372, 70700-0174, 70700-0175, 70700-0176, 00075-8003, 00075-8004, 00703-5720, 00703-5730, 16714-0465, 16714-0500, 16729-0120, 16729-0228, 39822-2120, 39822-2180, 39822-2200, 42367-0121, 45963-0781, 45963-0790, 50742-0428, 00069-9144, 00409-0369, 67457-0531, 67457-0532, 69097-0369, 69097-0371, 57884-3021 |
| Etoposide | HCPCS | C9414, C9425, J8560, J9181, J9182 |
| | NDC | 00378-3266, 00703-5653, 55390-0291, 55390-0292, 55390-0293, 55390-0491, 55390-0492, 55390-0493, 63323-0104, 00703-5657, 00703-5656, 16729-0114, 68001-0265, 00015-3404, 16729-0262 |
| Gemcitabine | HCPCS | J9201, J9198 |
| | NDC | 68001-0359, 68001-0350, 68001-0348, 68001-0342, 16714-0909, 16714-0930, 16729-0391, 16729-0419, 16729-0423, 00703-5775, 00703-5778, 00002-7502, 00781-3282, 00781-3283, 16729-0092, 16729-0117, 16729-0118, 55390-0391, 67457-0462, 67457-0463, 67457-0464, 68001-0282, 68083-0148, 68083-0149, 69097-0313, 69097-0314, 00409-0181, 00409-0182, 00409-0183, 00409-0185, 00409-0187, 25021-0209, 25021-0234, 25021-0235, 42236-0001, 42236-0002, 55111-0686, 55111-0687, 70860-0204, 70860-0205, 63323-0102, 63759-3028, 63759-3029, 71288-0117, 72485-0221, 72485-0222, 72485-0223, 00002-7501, 00143-9394, 00143-9395, 00409-0186, 23155-0213, 23155-0528, 25021-0208, 45963-0619, 63323-0125, 63323-0126, 67457-0616, 67457-0617, 67457-0618, 00069-3857, 00069-3858, 00069-3859, 00591-3562, 00591-3563, 23155-0214, 23155-0483, 23155-0484, 23155-0529, 25021-0239, 45963-0612, 45963-0620, 45963-0623, 45963-0624, 45963-0636, 47335-0153, 47335-0154, 50742-0496, 50742-0497, 50742-0498, 16729-0426, 62756-0008, 62756-0073, 62756-0102, 62756-0219, 62756-0321, 62756-0438, 62756-0533, 62756-0614, 62756-0746, 62756-0974, 71288-0113, 71288-0114, 60505-6113, 60505-6114, 60505-6115 |
| Irinotecan | HCPCS | C9474, J9206 |
| | NDC | 66758-0048, 68001-0284, 69171-0398, 68001-0480, 68083-0381, 70700-0169, 45963-0614, 55150-0352, 55150-0353, 55150-0354, 68001-0425, 68001-0426, 72485-0213, 68083-0382, 70700-0170, 00009-1111, 00009-7529, 00143-9583, 00143-9701, 00143-9702, 00703-4432, 00703-4434, 15054-0043, 16714-0027, 16714-0131, 25021-0214, 25021-0230, 47335-0937, 47335-0953, 50742-0401, 50742-0402, 59923-0702, 59923-0714, 59923-0715, 59923-0716, 61703-0349, 63323-0193, 72485-0211, 72485-0212, 16714-0725, 16714-0726, 23155-0179 |
| Lomustine | HCPCS | S0178 |
| | NDC | 00015-3030, 00015-3031, 00015-3032, 58181-3030, 58181-3031, 58181-3032, 58181-3040, 58181-3041, 58181-3042, 58181-3043 |
| Lurbinectedin | HCPCS | J9223 |
| | NDC | 68727-0712, |
| Paclitaxel | HCPCS | C9127, C9431, J9264, J9265, J9267 |
| | NDC | 47781-0595, 55390-0114, 55390-0304, 55390-0314, 66758-0043, 67457-0434, 67457-0449, 67457-0471, 68083-0178, 68083-0179, 68083-0180, 68817-0134, 70860-0200, 00703-3216, 00703-3217, 00703-3213, 00703-3218, 16714-0137, 69539-0158, 69539-0159, 69539-0157, 72205-0063, 72205-0062, 72205-0061, 00703-4764, 00703-4768, 44567-0504, 44567-0505, 44567-0506, 45963-0613, 61703-0342, 63323-0763, 00069-0076, 00069-0078, 00069-0079, 00703-4766, 00703-4767, 25021-0213, 68001-0516, 47781-0593, 47781-0594, 51991-0937, 51991-0938, 70860-0215 |
| Pemetrexed | HCPCS | C9213, J9304, J9305 |
| | NDC | 00002-7623, 00002-7640, 67184-0503 |
| Temozolomide | HCPCS | C1086, C9253, J8700, J9328 |
| | NDC | 54868-4142, 54868-5348, 54868-5350, 54868-5354, 54868-5980, 62175-0240, 62175-0241, 62175-0242, 62175- |

| | | |
|---|---|---|
| | | 0243, 62175-0244, 62175-0245, 64144-0501, 64144-0502, 64144-0503, 64144-0504, 64144-0505, 64144-0506, 64980-0333, 64980-0334, 64980-0335, 64980-0336, 64980-0337, 64980-0338, 65162-0801, 65162-0802, 65162-0803, 65162-0804, 65162-0805, 65162-0806, 67877-0537, 67877-0538, 67877-0539, 67877-0540, 67877-0541, 67877-0542, 69189-7638, 00085-0381, 16729-0048, 16729-0050, 16729-0051, 16729-0129, 16729-0130, 40051-0604, 40051-0605, 40051-0606, 40051-0607, 40051-0608, 40051-0609, 47335-0893, 00054-0320, 00054-0321, 00054-0322, 00054-0323, 00054-0324, 00054-0325, 16729-0049, 50268-0761, 50268-0762, 47335-0890, 47335-0891, 47335-0892, 47335-0929, 47335-0930, 62559-0921, 62559-0920, 62559-0922, 62559-0923, 62559-0924, 62559-0925, 00085-3004, 00085-1366, 00085-1381, 00085-1417, 00085-1425, 00085-1430, 00085-1519, 00093-7599, 00093-7600, 00093-7601, 00093-7602, 00093-7638, 00093-7639, 00378-5260, 00378-5261, 00378-5262, 00378-5263, 00378-5264, 00378-5265, 00527-1777, 00527-1778, 00527-1779, 00527-1780, 00527-1781, 00527-1782, 00781-2691, 00781-2692, 00781-2693, 00781-2694, 00781-2695, 00781-2696, 42737-0101, 42737-0102, 42737-0103, 42737-0104, 42737-0105, 42737-0106, 43975-0252, 43975-0253, 43975-0254, 43975-0255, 43975-0257, 50268-0763, 51862-0083, 51862-0084, 51862-0085, 51862-0086, 51862-0087, 51862-0088, 75834-0132, 75834-0142, 75834-0143, 75834-0144, 75834-0145, 43975-0256, 59923-0703, 59923-0704, 59923-0705, 59923-0706, 59923-0707, 59923-0708, 59923-0709, 59923-0710, 59923-0711, 59923-0712, 59923-0713, |
| Topotecan | HCPCS | J8705, J9350, J9351 |
| | NDC | 66758-0051, 66435-0410, 67457-0474, 00007-4201, 00007-4205, 00007-4207, 00069-0075, 00078-0672, 00078-0673, 00078-0674, 00409-0302, 00703-4714, 16729-0151, 16729-0243, 45963-0615, 55390-0370, 62756-0023, 63323-0762, 67457-0662, 71288-0127, 25021-0206, 25021-0236, 25021-0824, 50742-0404 |
| Vinorelbine | HCPCS | C9440, J9390 |
| | NDC | 55390-0069, 55390-0070, 00008-0045, 64370-0532, 61703-0341, 00069-0099, 00069-0103, 00069-0205, 00703-4182, 00703-4183, 25021-0204, 45963-0607, 66758-0045, 67457-0431, 67457-0479, 67457-0481, 67457-0482, 50742-0420, 50742-0427 |

**eTable 3. Characteristics of melanoma cases in SEER-Medicare by atopic disease history among all melanoma cases**

| Characteristic | No history of atopy (N=19,408 (65%)) | History of atopy (N=10,548 (35%)) | p-value[a] |
|---|---|---|---|
| **Male (%)** | 12,156 (63%) | 5,761 (55%) | **<0.001** |
| **Median age at diagnosis (IQR)** | 73 (68, 80) | 79 (74, 85) | **<0.001** |
| **Age group** | | | **<0.001** |
| 65-69 | 6,073 (31%) | 942 (8.9%) | |
| 70-74 | 4,611 (24%) | 2,122 (20%) | |
| 75-79 | 3,355 (17%) | 2,427 (23%) | |
| 80-84 | 2,574 (13%) | 2,354 (22%) | |
| 85+ | 2,795 (14%) | 2,703 (26%) | |
| **Race/Ethnicity** | | | 0.377 |
| Non-Hispanic White | 18,915 (97%) | 10,265 (97%) | |
| Non-Hispanic Black | 64 (0.3%) | 48 (0.5%) | |
| Non-Hispanic Other | 101 (0.5%) | 59 (0.6%) | |
| Hispanic | 328 (1.7%) | 176 (1.7%) | |
| **Comorbidity Score** | | | **<0.001** |
| 0 | 11,671 (60%) | 4,556 (43%) | |
| 1 | 2,499 (13%) | 2,145 (20%) | |
| 2 | 2,878 (15%) | 1,778 (17%) | |
| 3+ | 2,360 (12%) | 2,069 (20%) | |
| **Stage at diagnosis** | | | **<0.001** |
| Localized | 15,996 (82%) | 8,910 (84%) | |
| Distant site involved | 1,293 (6.7%) | 555 (5.3%) | |
| Regional | 2,119 (11%) | 1,083 (10%) | |

[a] p-value corresponding to either log-rank test for median survival, two-tailed t-test for age at diagnosis, or □2 test of independence for all others.

**eTable 4. Hazard ratios and p-values from univariate analyses of demographic and prognostic factors for overall survival in all melanoma cases and cumulative incidence of brain metastatsis in individuals with non-metastatic disease at diagnosis**

| Characteristic | Overall survival | | Cumulative incidence of brain metastasis | |
|---|---|---|---|---|
| | Univariate HR (95% CI) | P value | Univariate HR (95% CI) | P value |
| **Sex** | | | | |
| Male | 1.0 | ref | 1.0 | ref |
| Female | 0.86 (0.82-0.89) | <0.001 | 0.69 (0.62-0.76) | <0.001 |
| **Age at diagnosis** | 1.10 (1.10-1.10) | <0.001 | 1.00 (1.00-1.01) | 0.232 |
| **Race/Ethnicity** | | | | |
| Non-Hispanic White | 1.0 | ref | 1.0 | ref |
| Non-Hispanic Black | 2.09 (1.63-2.69) | <0.001 | 4.08 (2.65-6.27) | <0.001 |
| Non-Hispanic Other | 1.49 (1.17-1.89) | <0.001 | 2.14 (1.32-3.45) | 0.002 |
| Hispanic | 1.13 (0.98-1.31) | 0.099 | 1.70 (1.27-2.28) | <0.001 |
| **Stage at diagnosis** | | | | |
| Localized | 1.0 | ref | 1.0 | ref |
| Regional | 9.36 (8.84-9.90) | <0.001 | 26.72 (23.70-30.13) | <0.001 |
| Distant site involved | 2.72 (2.58-2.86) | <0.001 | 6.66 (5.95-7.46) | <0.001 |
| **Tumor size / AJCC T staging** | | | | |
| < 1.0 mm (T1) | 1.0 | ref | 1.0 | ref |
| >1-2 mm (T2) | 1.58 (1.47-1.71) | <0.001 | 4.09 (3.30-5.08) | <0.001 |
| >2-4 mm (T3) | 2.63 (2.44-2.83) | <0.001 | 9.37 (7.69-11.41) | <0.001 |
| >4 mm (T4) | 4.52 (4.21-4.85) | <0.001 | 14.64 (12.05-17.78) | <0.001 |
| Unknown | 2.48 (2.36-2.60) | <0.001 | 6.61 (5.61-7.80) | <0.001 |
| **Surgical Size** | | | | |
| <3.1 cm | 1.0 | ref | 1.0 | ref |
| >3.0 cm | 1.56 (1.49-1.64) | <0.001 | 1.22 (1.08-1.38) | 0.001 |
| None/Unknown | 2.62 (2.51-2.75) | <0.001 | 2.17 (1.94-2.42) | <0.001 |
| **Primary Site** | | | | |
| Upper limb and shoulder | ref | | | |
| Head and neck | 1.38 (1.31-1.46) | <0.001 | 1.61 (1.40-1.85) | <0.001 |
| Lower limb and hip | 1.09 (1.02-1.16) | 0.011 | 1.32 (1.11-1.56) | 0.001 |
| Trunk | 0.99 (0.94-1.05) | 0.740 | 1.28 (1.11-1.49) | <0.001 |
| Other or site unspecified | 6.46 (6.01-6.95) | <0.001 | 12.96 (11.01-15.26) | <0.001 |
| **Breslow depth of invasion** | | | | |
| < 1.0 mm | 1.0 | ref | 1.0 | ref |
| >1-2 mm | 1.37 (1.28-1.48) | <0.001 | 2.55 (2.11-3.09) | <0.001 |
| >2-4 mm | 2.25 (2.09-2.41) | <0.001 | 6.16 (5.20-7.30) | <0.001 |
| >4 mm | 4.03 (3.78-4.30) | <0.001 | 9.99 (8.51-11.72) | <0.001 |
| Unknown | 2.44 (2.32-2.56) | <0.001 | 4.67 (4.06-5.37) | <0.001 |
| **Ulceration** | | | | |

| | | | | |
|---|---|---|---|---|
| Not present | 1.0 | ref | 1.0 | ref |
| Present | 2.98 (2.83-3.14) | <0.001 | 6.14 (5.43-6.94) | <0.001 |
| Unknown | 2.35 (2.25-2.45) | <0.001 | 3.53 (3.14-3.96) | <0.001 |
| **Received surgery for primary tumor (%)** | | | | |
| No | 1.0 | ref | 1.0 | ref |
| Yes | 0.38 (0.36-0.39) | <0.001 | 0.37 (0.33-0.41) | <0.001 |
| **Received radiation treatment (%)** | | | | |
| No | 1.0 | ref | 1.0 | ref |
| Yes | 3.76 (3.54-4.01) | <0.001 | 13.07 (11.75-14.54) | <0.001 |
| **Received chemotherapy (%)** | | | | |
| No | 1.0 | ref | 1.0 | ref |
| Yes | 1.62 (2.03-2.35) | <0.001 | 7.31 (5.89-7.54) | <0.001 |
| **Received checkpoint immunotherapy (%)** | | | | |
| No | 1.0 | ref | 1.0 | ref |
| Yes | 2.07 (1.94-2.20) | <0.001 | 9.74 (8.82-10.76) | <0.001 |
| **Comorbidity Score** | | | | |
| 0 | 1.0 | ref | 1.0 | ref |
| 1 | 1.91 (1.80-2.02) | <0.001 | 1.09 (0.95-1.25) | 0.214 |
| 2 | 1.74 (1.64-1.83) | <0.001 | 1.13 (0.99-1.29) | 0.064 |
| 3+ | 3.16 (3.01-3.33) | <0.001 | 1.19 (1.03-1.37) | 0.018 |
| **Any atopic condition** | | | | |
| No | 1.0 | ref | 1.0 | ref |
| Yes | 1.17 (1.13-1.22) | <0.001 | 0.79 (0.71-0.87) | <0.001 |
| **Allergic rhinitis** | | | | |
| No | 1.0 | ref | 1.0 | ref |
| Yes | 1.01 (0.96-1.07) | 0.578 | 0.84 (0.73-0.96) | 0.011 |
| **Atopic dermatitis** | | | | |
| No | 1.0 | ref | 1.0 | ref |
| Yes | 1.18 (1.13-1.24) | <0.001 | 0.71 (0.62-0.81) | <0.001 |
| **Asthma** | | | | |
| No | 1.0 | ref | 1.0 | ref |
| Yes | 1.27 (1.20-1.36) | <0.001 | 0.92 (0.78-1.09) | 0.334 |
| **Allergic/atopic conjunctivitis** | | | | |
| No | 1.0 | ref | 1.0 | ref |
| Yes | 0.94 (0.83-1.06) | 0.290 | 0.74 (0.53-1.02) | 0.065 |

**eTable 5. Hazard ratios and p-values from multivariable[a] subset and sensitivity analyses for overall survival in all melanoma cases and cumulative incidence of brain metastasis in individuals with non-metastatic disease at diagnosis[b]**

| Characteristic | Overall survival | | | | Cumulative incidence of brain metastasis | | | |
|---|---|---|---|---|---|---|---|---|
| | N | Deaths | Multivariable HR (95% CI) | P value | N | BrM events | Multivariable HR (95% CI) | P value |
| **Males only** | | | | | | | | |
| No history of atopic disease | 7003 | 2247 | 1.0 | Ref | 791 | 161 | 1.0 | Ref |
| History of atopic disease | 3372 | 1356 | 0.88 (0.82-0.94) | <0.001 | 311 | 51 | 0.96 (0.69-1.34) | 0.801 |
| No history of allergic rhinitis | 8739 | 2998 | 1.0 | Ref | 966 | 190 | 1.0 | Ref |
| History of allergic rhinitis | 1,636 | 605 | 0.88 (0.81-0.96) | 0.005 | 136 | 22 | 0.91 (0.58-1.44) | 0.702 |
| No history of atopic dermatitis | 8,577 | 2,834 | 1.0 | Ref | 930 | 188 | 1.0 | Ref |
| History of atopic dermatitis | 1,798 | 769 | 0.86 (0.80-0.94) | <0.001 | 172 | 24 | 0.84 (0.54-1.30) | 0.433 |
| No history of asthma | 9,584 | 3,275 | 1.0 | Ref | 1035 | 200 | 1.0 | Ref |
| History of asthma | 791 | 328 | 0.87 (0.78-0.98) | 0.024 | 67 | 12 | 1.10 (0.60-2.02) | 0.767 |
| No history of allergic/atopic conjunctivitis | 10173 | 3531 | 1.0 | Ref | 1087 | 209 | -- | -- |
| History of allergic/atopic conjunctivitis | 202 | 72 | 0.73 (0.58-0.93) | 0.010 | 15 | <11 | -- | -- |
| **Females only** | | | | | | | | |
| No history of atopic disease | 4185 | 1225 | 1.0 | Ref | 437 | 60 | 1.0 | Ref |
| History of atopic disease | 2721 | 923 | 0.87 (0.8-0.95) | 0.002 | 265 | 31 | 0.82 (0.52-1.30) | 0.395 |
| No history of allergic rhinitis | 5,532 | 1,711 | 1.0 | Ref | 570 | 77 | 1.0 | Ref |
| History of allergic rhinitis | 1,374 | 437 | 0.90 (0.81-1.01) | 0.062 | 132 | 14 | 0.66 (0.36-1.21) | 0.177 |
| No history of atopic dermatitis | 5,513 | 1674 | 1.0 | Ref | 568 | 75 | 1.0 | Ref |
| History of atopic dermatitis | 1,393 | 474 | 0.83 (0.75-0.92) | <0.001 | 134 | 16 | 0.90 (0.51-1.58) | 0.719 |
| No history of asthma | 6,106 | 1,840 | 1.0 | Ref | 630 | 84 | -- | -- |
| History of asthma | 800 | 308 | 1.07 (0.95-1.21) | 0.271 | 72 | <11 | -- | -- |
| No history of allergic/atopic conjunctivitis | 6620 | 2053 | 1.0 | Ref | 676 | 89 | -- | -- |
| History of allergic/atopic conjunctivitis | 286 | 95 | 0.92 (0.75-1.13) | 0.436 | 26 | <11 | -- | -- |
| **Regional disease only** | | | | | | | | |
| No history of atopic dermatitis | 344 | 203 | 1.0 | Ref | 302 | 46 | 1.0 | Ref |
| History of atopic dermatitis | 52 | 27 | 0.91 (0.77-1.07) | 0.252 | 47 | 11 | 0.85 (0.61-1.20) | 0.365 |
| **Distant spread without BrM at diagnosis** | | | | | | | | |
| No history of atopic dermatitis | 344 | 279 | 1.0 | Ref | 302 | 93 | 1.0 | Ref |
| History of atopic dermatitis | 52 | 39 | 0.64 (0.45-0.93) | 0.018 | 47 | 11 | 0.37 (0.18-0.75) | 0.006 |
| **Number of atopic conditions** | | | | | | | | |
| 0 | 11188 | 3472 | 1.0 | Ref | 1228 | 221 | 1.0 | Ref |
| 1 | 4316 | 1611 | 0.89 (0.84-0.94) | <0.001 | 427 | 68 | 1.03 (0.77-1.37) | 0.86 |
| 2+ | 1777 | 668 | 0.84 (0.78-0.92) | <0.001 | 149 | 14 | 0.56 (0.32-0.97) | 0.037 |

| | | | | | | | | |
|---|---|---|---|---|---|---|---|---|
| **Distant spread at diagnosis and treated with ICI[c]** | | | | | | | | |
| No history of atopic disease | 86 | 59 | 1.0 | Ref | 78 | 29 | 1.0 | ref |
| History of atopic disease | 35 | 21 | 0.31 (0.15-0.64) | 0.001 | 34 | 12 | 0.81 (0.30-2.15) | 0.668 |
| No history of atopic dermatitis | 108 | 73 | -- | -- | 100 | 36 | -- | -- |
| History of atopic dermatitis | 13 | <11 | -- | -- | 12 | <11 | -- | -- |
| **Distant spread at diagnosis and not treated with ICI** | | | | | | | | |
| No history of atopic disease | 88 | 72 | 1.0 | Ref | 73 | 18 | -- | -- |
| History of atopic disease | 48 | 39 | 1.41 (0.87-2.27) | 0.165 | 43 | <11 | -- | -- |
| No history of atopic dermatitis | 114 | 94 | 1.0 | Ref | 96 | 24 | -- | -- |
| History of atopic dermatitis | 22 | 17 | 0.95 (0.48-1.87) | 0.879 | 20 | <11 | -- | -- |

[a] All Cox proportional hazards models adjusted for age at diagnosis, sex, race/ethnicity, comorbidity score, stage at diagnosis, primary site, tumor size, depth of invasion, presence of ulceration, and treatment receipt (including resection, radiation, and chemotherapy).

[b] Cells were suppressed if their value was less <11, or if that cell's value would allow deriving a suppressed cell's value.

[c] 2012-2017 only.

**eFigure 1. Flowchart of inclusion/exclusion criteria**

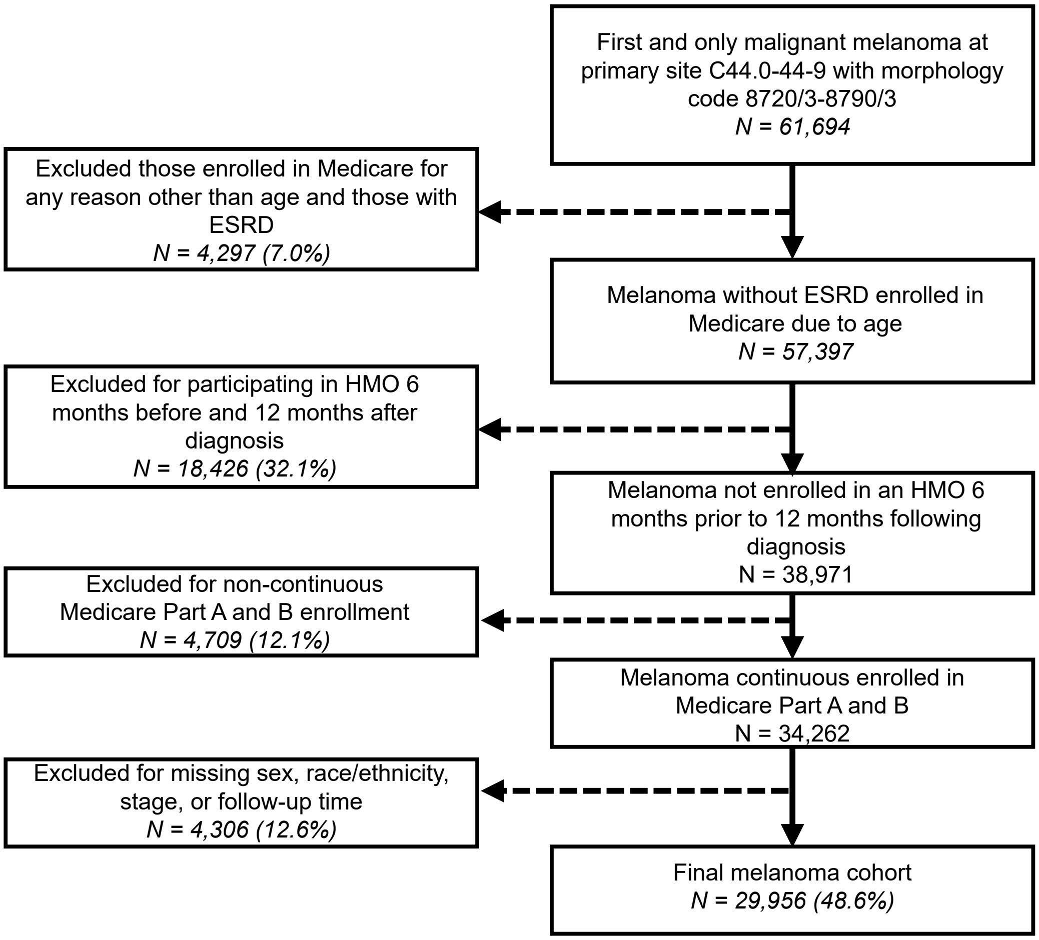

**eFigure 2. Correlation between selected prognostic factors among SEER-Medicare melanoma cases**

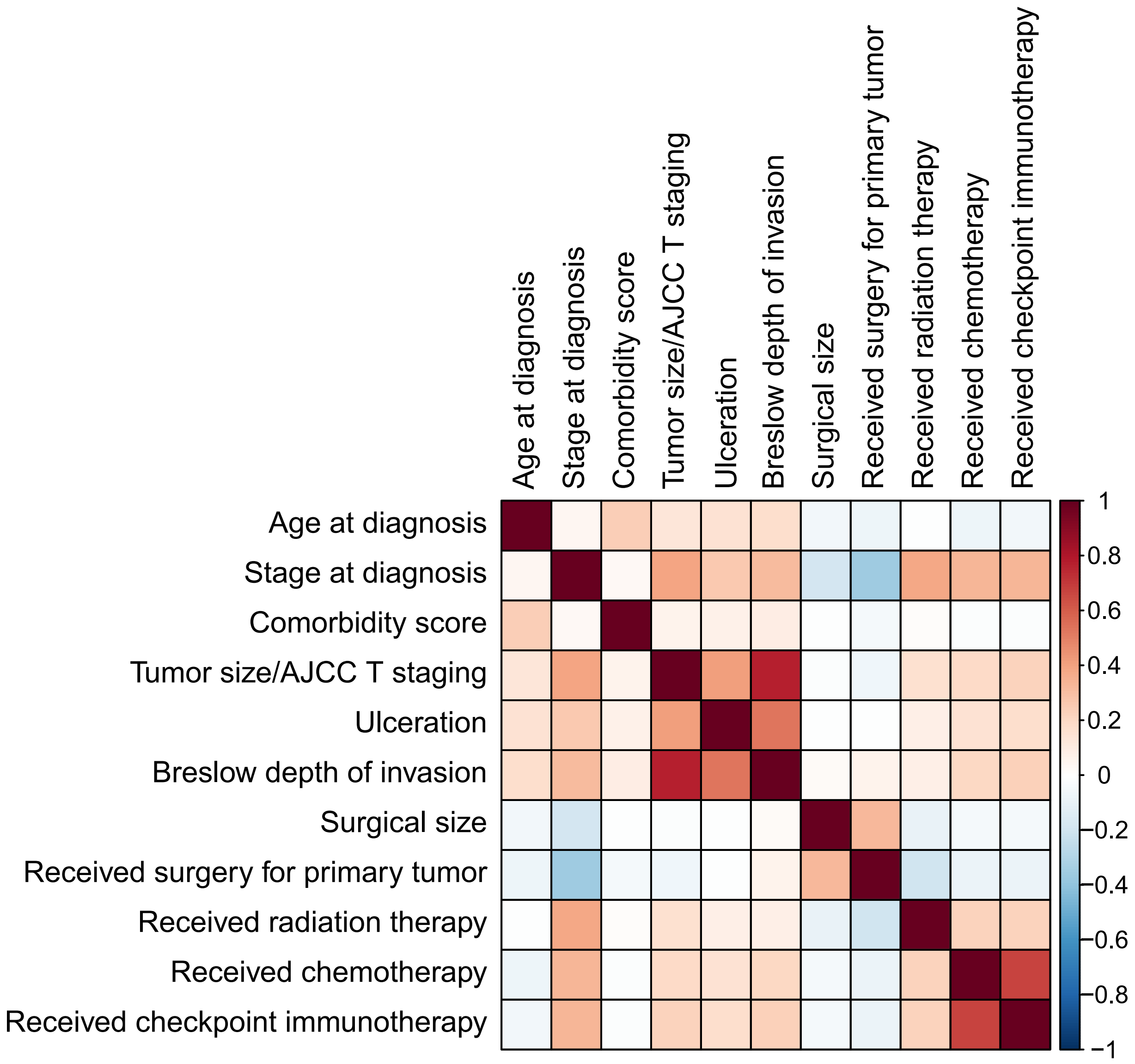

**eFigure 3. Frequency of atopic conditions at time of melanoma diagnosis among those with brain metastasis by metastasis timing and sex**

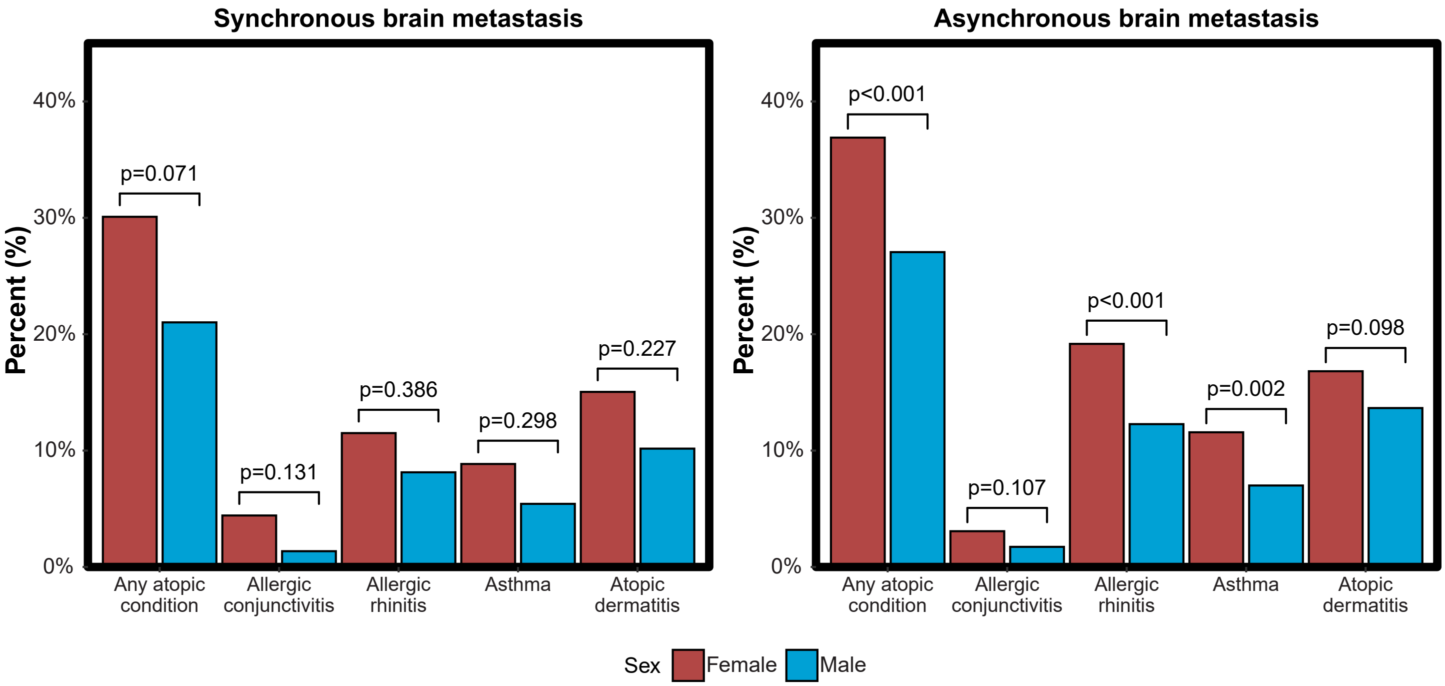

**eFigure 4. Forest plots containing adjusted[a] odds ratios (OR) with 95%CI and FDR-adjusted p-values confidence of receiving treatment for melanoma.**

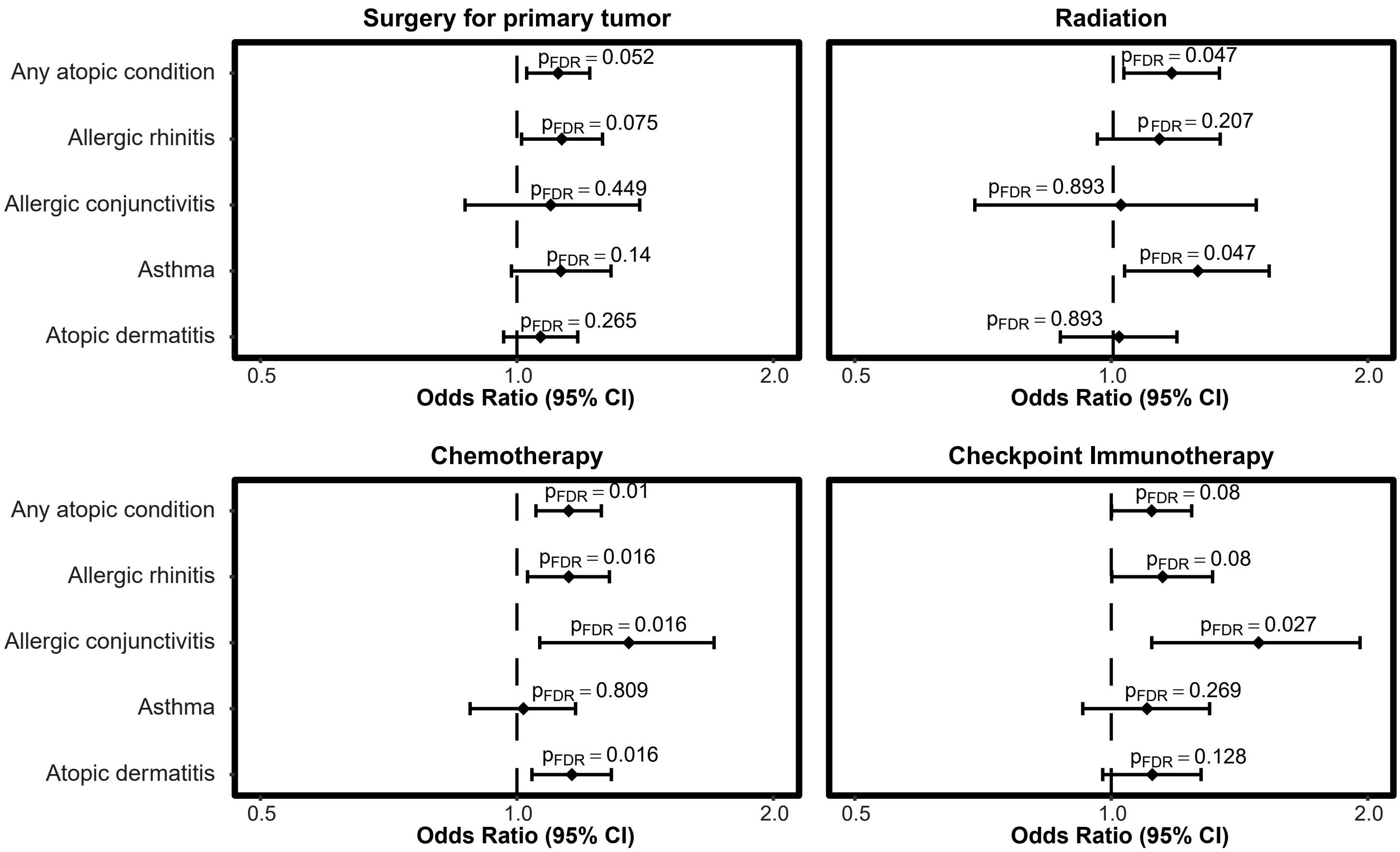


a. All logistic regression models adjusted for age at diagnosis, sex, race/ethnicity, comorbidity score, stage at diagnosis, primary site, tumor size, depth of invasion, and presence of ulceration. Radiation and chemotherapy were additionally adjusted for surgery for primary tumor.

**eFigure 5. Proportion of melanoma cases with brain metastases in melanoma by race/ethnicity A) overall, B) in males only, and C) in females only**

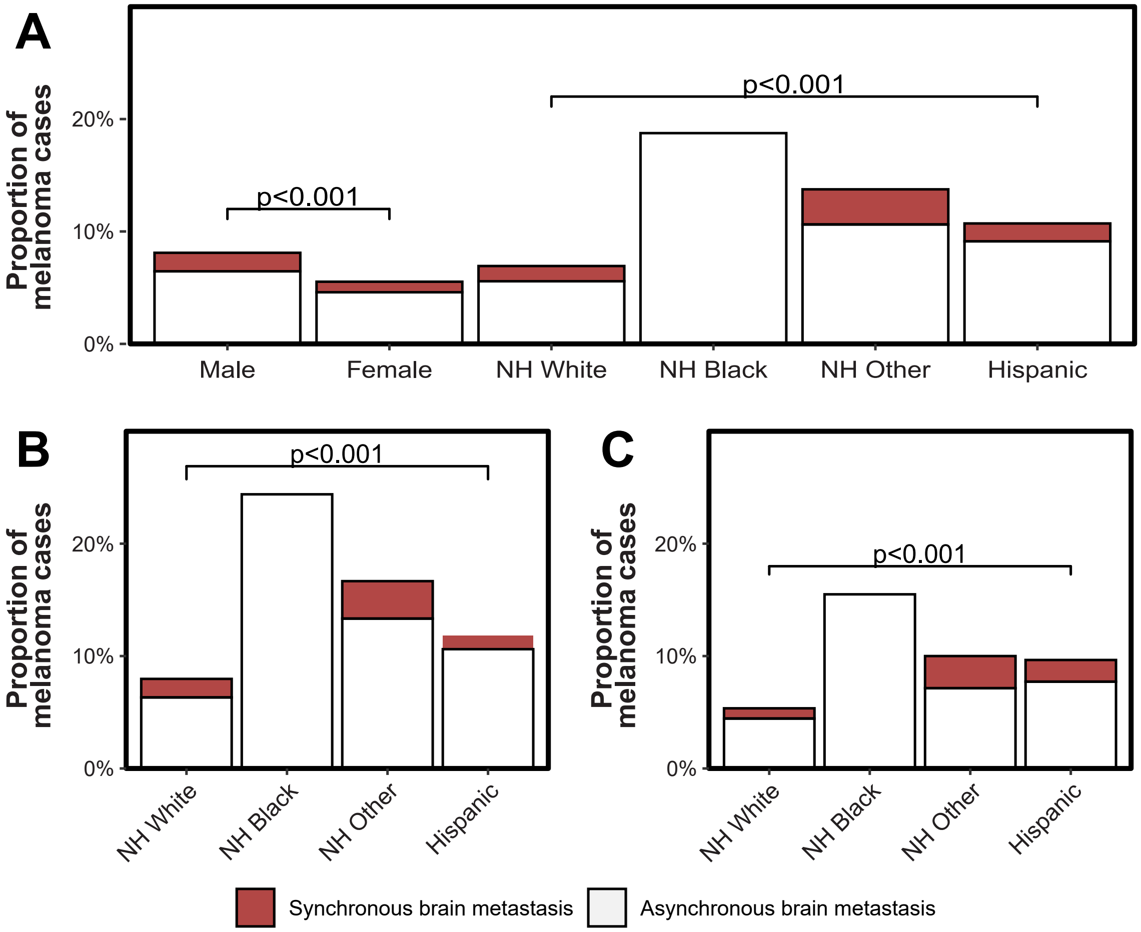

**eFigure 6. Overall and sex-specific hazard ratios[a] of death, 95% CI, and p-values[b] for individuals with melanoma by age of diagnosis for any atopic disorder and atopic dermatitis only**

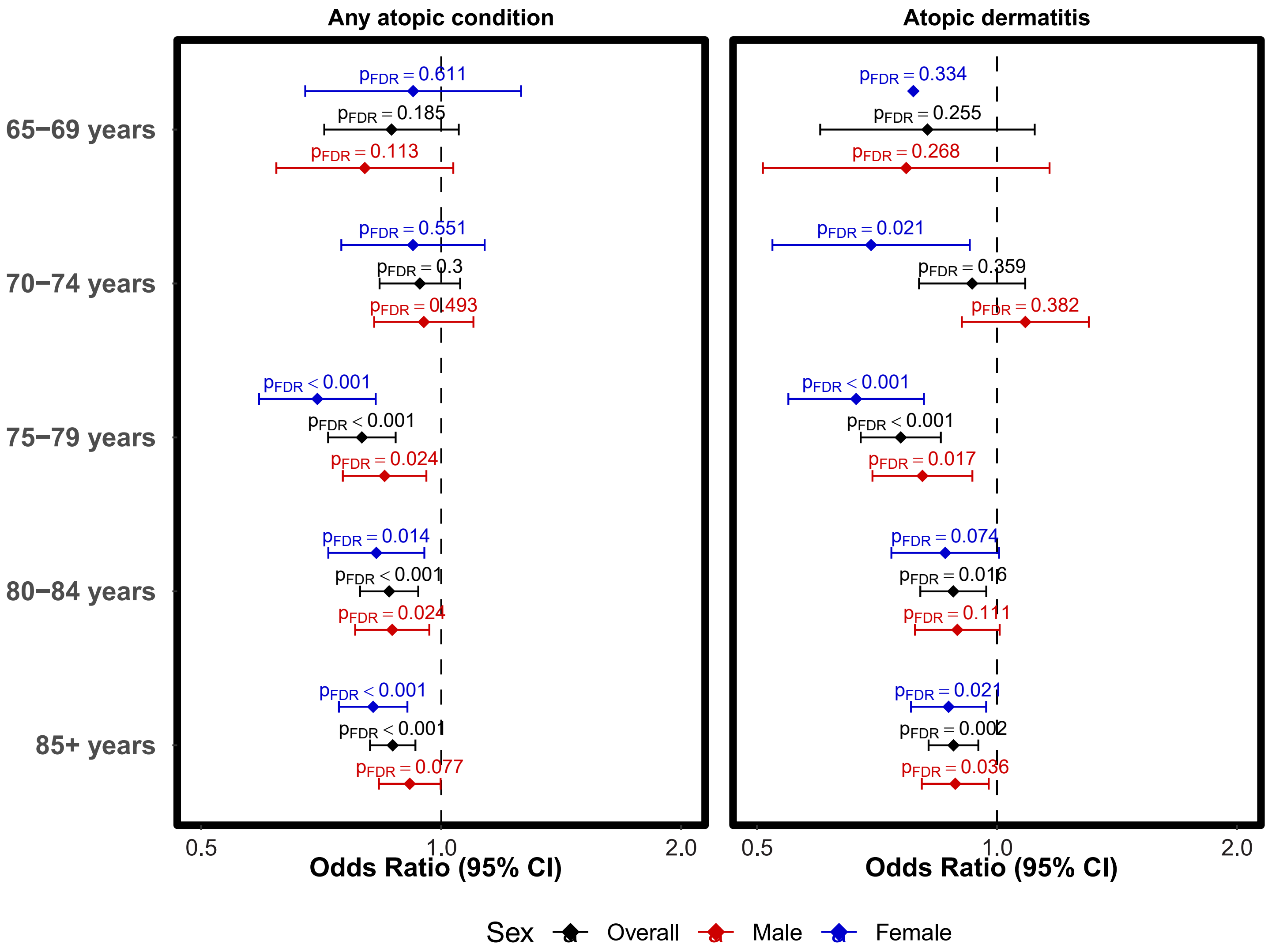


a. All Cox proportional hazards models adjusted for age at diagnosis, sex, race/ethnicity, comorbidity score, stage at diagnosis, primary site, tumor size, depth of invasion, presence of ulceration, and treatment receipt (including resection, radiation, and chemotherapy)

**eFigure 7. Adjusted[a] probability of survival in individuals with melanoma that had BrM at diagnosis or developed BrM by prior history of A) any atopic condition, and B) corresponding hazard ratios, 95% CIs and p-values[b], and adjusted probability of survival by prior history of C) allergic rhinitis, D) allergic/atopic conjunctivitis, E) asthma, and F) atopic dermatitis**

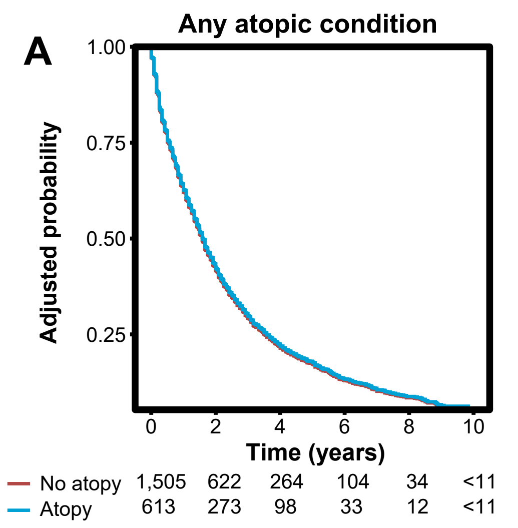


B

| Atopic condition | HR (95% CI) | P | $P_{FDR}$ |
|---|---|---|---|
| Any atopic condition | 0.98 (0.87−1.09) | 0.652 | 0.814 |
| Allergic rhinitis | 1.00 (0.87−1.15) | 0.983 | 0.983 |
| Allergic conjunctivitis | 0.87 (0.63−1.20) | 0.389 | 0.681 |
| Asthma | 0.93 (0.78−1.11) | 0.409 | 0.681 |
| Atopic dermatitis | 0.93 (0.81−1.07) | 0.314 | 0.681 |

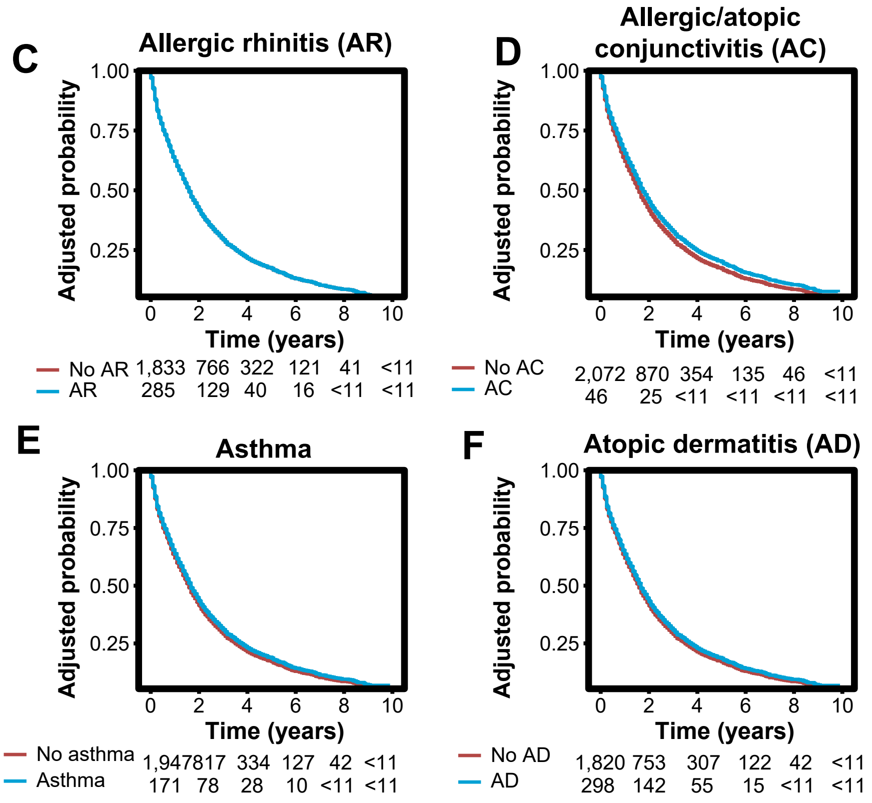


a. Adjusted for age at diagnosis, sex, race/ethnicity, comorbidity score, stage at diagnosis, primary site, tumor size, depth of invasion, presence of ulceration, and treatment receipt (including resection, radiation, and chemotherapy)
b. Number of individuals at risk is suppressed when <11 per SEER-Medicare Data Use Restrictions

**eFigure 8. Adjusted[a] probability of survival in individuals with melanoma that had BrM at diagnosis (synchronous BrM) by prior history of A) any atopic condition, and B) corresponding hazard ratios, 95% CIs and p-values[b], and adjusted probability of survival by prior history of C) allergic rhinitis, D) allergic/atopic conjunctivitis, E) asthma, and F) atopic dermatitis**

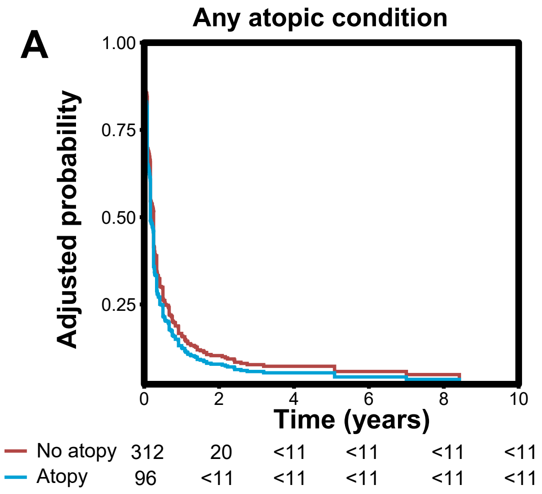


B

| Atopic condition | HR (95% CI) | P | $P_{FDR}$ |
|---|---|---|---|
| Any atopic condition | 1.21 (0.93−1.58) | 0.164 | 0.460 |
| Allergic rhinitis | 1.23 (0.85−1.79) | 0.278 | 0.463 |
| Allergic conjunctivitis | 1.15 (0.58−2.29) | 0.689 | 0.861 |
| Asthma | 1.32 (0.88−2.00) | 0.184 | 0.460 |
| Atopic dermatitis | 0.97 (0.69−1.37) | 0.872 | 0.872 |

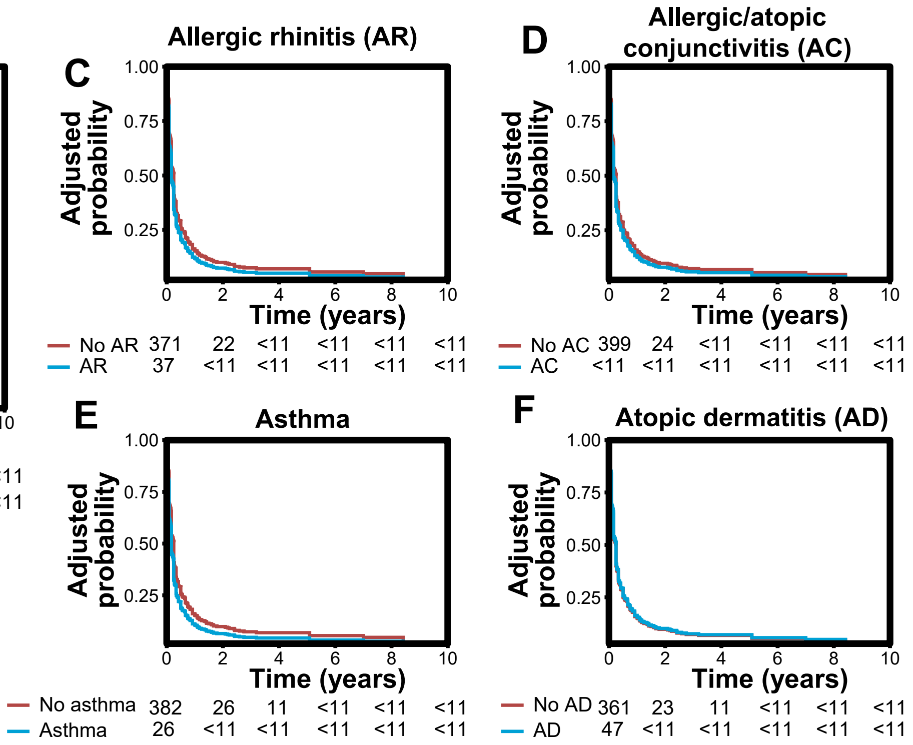


a. Adjusted for age at diagnosis, sex, race/ethnicity, comorbidity score, stage at diagnosis, primary site, tumor size, depth of invasion, presence of ulceration, and treatment receipt (including resection, radiation, and chemotherapy)
b. Number of individuals at risk is suppressed when <11 per SEER-Medicare Data Use Restrictions

**eFigure 9. Adjusted[a] probability of survival in individuals with melanoma that developed BrM (asynchronous BrM) by prior history of A) any atopic condition, and B) corresponding hazard ratios, 95% CIs and p-values[b], and adjusted probability of survival by prior history of C) allergic rhinitis, D) allergic/atopic conjunctivitis, E) asthma, and F) atopic dermatitis**

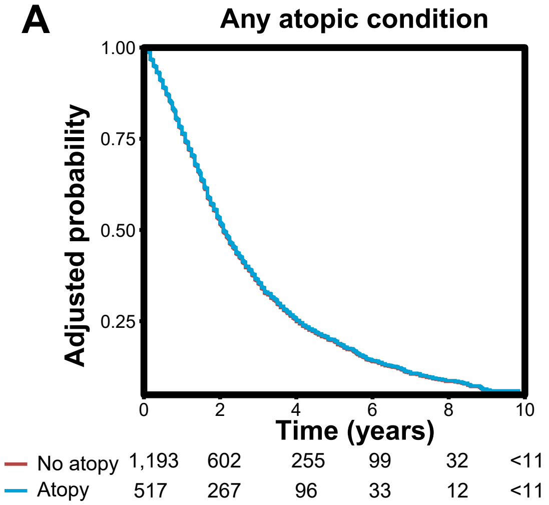


B

| Atopic condition | HR (95% CI) | P | $P_{FDR}$ |
|---|---|---|---|
| Any atopic condition | 0.99 (0.87−1.12) | 0.842 | 0.842 |
| Allergic rhinitis | 1.06 (0.91−1.24) | 0.455 | 0.569 |
| Allergic conjunctivitis | 0.86 (0.59−1.24) | 0.408 | 0.569 |
| Asthma | 0.90 (0.73−1.09) | 0.277 | 0.569 |
| Atopic dermatitis | 0.93 (0.80−1.09) | 0.385 | 0.569 |

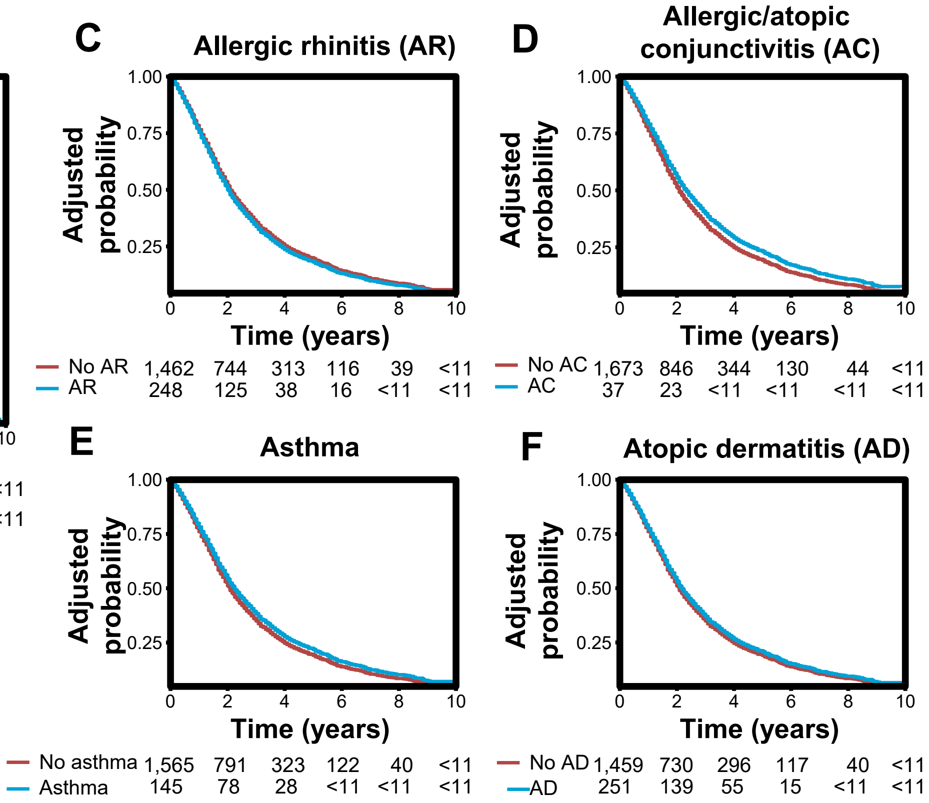


a. Adjusted for age at diagnosis, sex, race/ethnicity, comorbidity score, stage at diagnosis, primary site, tumor size, depth of invasion, presence of ulceration, and treatment receipt (including resection, radiation, and chemotherapy)
b. Number of individuals at risk is suppressed when <11 per SEER-Medicare Data Use Restrictions

**eFigure 10. Adjusted[a] probability of BrM-free survival in melanoma by prior history of A) any atopic condition, and B) corresponding hazard ratios, 95% CIs and p-values, and adjusted probability of survival by prior history of C) allergic rhinitis, D) allergic/atopic conjunctivitis, E) asthma, and F) atopic dermatitis**

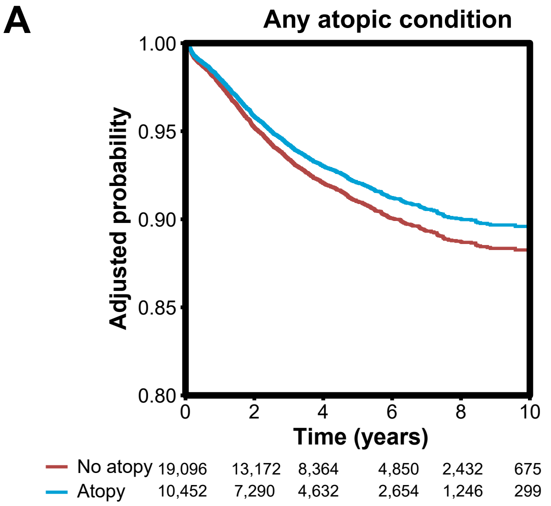


B

| Atopic condition | HR (95% CI) | P | $P_{FDR}$ |
|---|---|---|---|
| Any atopic condition | 0.85 (0.76−0.94) | 0.003 | 0.006 |
| Allergic rhinitis | 0.94 (0.82−1.08) | 0.356 | 0.445 |
| Allergic conjunctivitis | 0.79 (0.57−1.10) | 0.157 | 0.262 |
| Asthma | 0.95 (0.80−1.13) | 0.551 | 0.551 |
| Atopic dermatitis | 0.75 (0.65−0.86) | <0.001 | <0.001 |

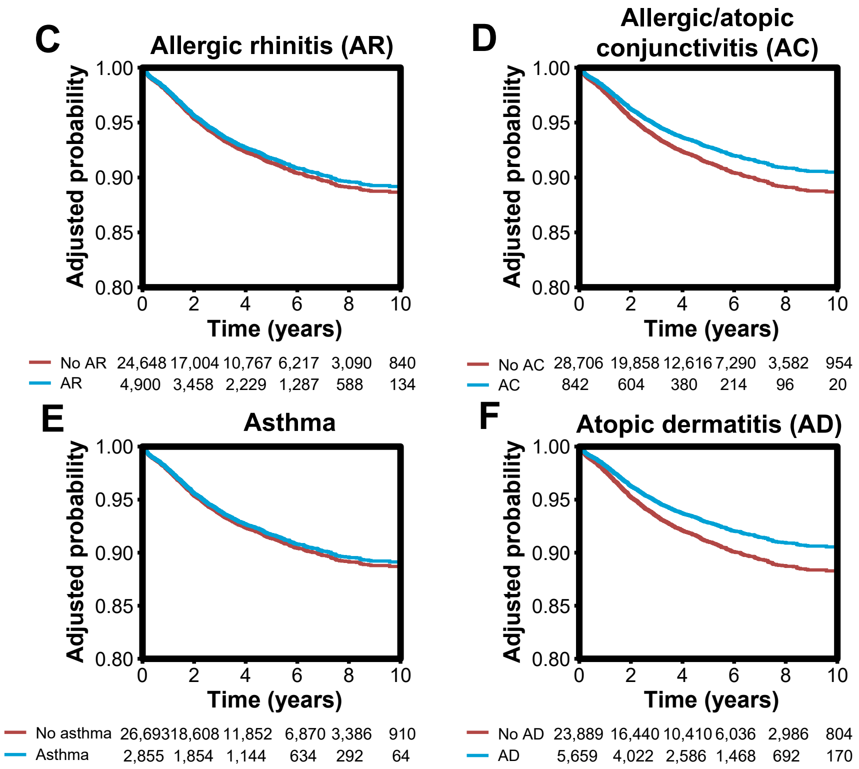


a. Adjusted for age at diagnosis, sex, race/ethnicity, comorbidity score, stage at diagnosis, primary site, tumor size, depth of invasion, presence of ulceration, and treatment receipt (including resection, radiation, and chemotherapy)
